# A Randomized, Double-Blind, Controlled Trial to Assess the Effects of Lactoferrin at Two Doses vs. Active Control on Immunological and Safety Parameters in Healthy Adults

**DOI:** 10.1101/2024.07.17.24310517

**Authors:** Ross D. Peterson, Liana L. Guarneiri, Caryn G. Adams, Meredith L. Wilcox, Anthony J. Clark, Nathan P. Rudemiller, Kevin C. Maki, Carrie-Anne Malinczak

## Abstract

**Background:** Recombinant human lactoferrin (rhLF) is of commercial interest for support of the immune system and iron homeostasis but is not currently available as a food ingredient.

**Objective:** The objective was to evaluate the immunogenicity/alloimmunization potential of Helaina rhLF (effera™) from *K. phaffii* over a 28-day period compared to bovine LF (bLF). It was hypothesized that rhLF would have an equal or lower immunogenic potential compared to bLF, which is permitted for use in conventional foods and infant formula in the EU and US.

**Methods:** Study 1 was a randomized, double-blind, parallel arm, controlled trial where 66 healthy adults were randomly allocated to 1 of 3 groups: high-dose rhLF (3.4 g/d), low-dose rhLF (0.34 g/d), or bLF (3.4 g/d) (clinicaltrials.gov: NCT06012669). Participants completed a 28-day (Day 28) supplementation period followed by a 28-day washout period (Day 56) and an additional 28-day follow-up period (Day 84). Study 2 was a 12-week observational study with no intervention that enrolled 24 healthy adults. In both studies, fasting blood was obtained on Days 0, 28, 56, and 84 for analysis of anti-bLF and anti-hLF antibody levels as the primary endpoint as well as other secondary safety endpoints including complete blood count, iron biomarkers, and metabolic panel.

**Results:** In Study 1, the change from baseline to Day 56 in serum anti-bLF antibodies in the bLF group (least squares geometric mean and 95% confidence interval for the post/pre ratio: 3.01; 2.08, 4.35) was greater than the changes in serum anti-hLF antibodies in the low-dose rhLF (1.07; 0.77, 1.49; P<0.001) and high-dose rhLF (1.02; 0.62, 1.70; P<0.001) groups. In Study 2, the post/pre ratio at Day 56 in serum anti-hLF and anti-bLF antibodies were 1.00 (0.93, 1.08) and 1.01 (0.94, 1.09), respectively, confirming no change in anti-hLF antibodies in Study 1 or Study 2. Changes in all safety outcomes in Study 1 were similar between groups and within normal ranges.

**Conclusion:** These results support the safety and tolerability of effera^TM^ as a food ingredient at an intake level up to 3.4 g/d in healthy adults.

## INTRODUCTION

Lactoferrin, an iron-binding glycoprotein, is a component of the whey fraction of milk from humans and other mammals (1). It is produced in the body by exocrine glands and neutrophils; thus, it is found in human milk, tears, saliva, gastrointestinal fluids, cerebrospinal fluid, and cells of the immune system (1, 2). Lactoferrin is consumed throughout the lifespan from different sources. Human milk is particularly rich in lactoferrin, containing 5-7 mg/mL in colostrum to 1-3 mg/mL in mature milk (3). After weaning, bovine milk lactoferrin (bLF) is commonly consumed through ingestion of cow’s milk and dairy products (4). Interest in supplementation with lactoferrin has peaked due to a growing body of research highlighting the physiological importance of lactoferrin for iron homeostasis and immune function (5, 6).

Approximately 40% of pregnant women and 32.5% of non-pregnant women globally had anemia (mostly due to iron deficiency) in 2016, highlighting the crucial need for iron supplementation (7). However, the bioavailability of oral non-heme iron supplements is highly variable and often poor (1-10% absorbed), and the high-dose iron supplements that are frequently administered often result in undesirable gastrointestinal side effects (8, 9). Although the mechanism is not fully understood, emerging evidence suggests that lactoferrin enhances intestinal absorption of iron and improves hemoglobin production (8, 10). A meta-analysis of 11 studies (1,262 participants) indicated that serum iron, serum ferritin, and hemoglobin showed greater improvements following daily supplementation with oral lactoferrin vs. oral ferrous sulfate, but fractional iron absorption was superior with ferrous sulfate vs. lactoferrin (10). The lactoferrin utilized for the studies included in this meta-analysis were bLF or recombinant human lactoferrin (rhLF) from transgenic rice. Regarding the immune system, Berthon, et al. recently conducted a systematic review and meta-analysis investigating the impact of lactoferrin supplementation on inflammation and immune function in humans (11). Eight out of 13 studies reported improvements in at least one biomarker of immune function, and 6 of 8 studies reported improvements in biomarkers of immune function. All the trials included in this meta-analysis involved bLF in doses ranging from 32.4 mg/d to 3 g/d, and all trials except one were conducted in adults.

Bovine milk lactoferrin is generally recognized as safe (GRAS) in the United States for use in term infant formula, sports foods, functional foods, chewing gum, and as an antimicrobial agent (12–16). Recombinant human lactoferrin was the subject of three GRAS Notices, but the Food and Drug Administration ceased to evaluate these notices at the request of the notifier (17–19). Despite this regulatory history, a comprehensive safety review recently compiled all in vivo studies including animal toxicology studies and human studies conducted on the hLFs from transgenic rice, cows and fungi (i.e., *Aspergillus niger*) (20). Overall, these studies support the general tolerance and safety of rhLF, however, a key safety question related to the immunogenicity/alloimmunization potential of rhLF was never evaluated (20). The question was raised by an Expert Panel at the Toxicology Forum in 2008 and another Expert Panel in 2023 and is related to the exogenous nature of rhLF (21). In this context, alloimmunization is the breakdown of tolerance to a self-protein, hLF, with the potential to elicit an immune response leading to adverse events (AEs). Potential AEs include autoimmunity and/or a targeted response against the endogenous protein, which would limit the inherent physiological properties of hLF. An established method to evaluate alloimmunization is testing for the development of treatment-emergent antibodies against the endogenous protein (22). Although rhLF has the same amino acid sequence as endogenous hLF present throughout the human body, there are differences in post-translational modifications due to the expression system of the recombinant form (23) and the potential to generate novel epitopes. Thus, the alloimmunization potential of rhLF taken as a food ingredient is warranted.

Helaina recombinant human lactoferrin (effera™) from *Komagataella phaffii* has been developed as a potential food ingredient and dietary supplement (23). Analytical analyses of the primary, secondary, and tertiary structure of Helaina rhLF compared to human milk LF and bLF were recently conducted (23). Recent preclinical safety studies demonstrated that Helaina rhLF has low allergenic risk potential, and it was well tolerated in rats at levels up to 2000 mg/kg/day (57x the intended commercial use) (24, 25).

The preclinical data suggest that the Helaina rhLF is safe for human exposure, allowing for the alloimmunization potential to be tested in humans. Two clinical studies were conducted simultaneously. The purpose of Study 1 was to evaluate the impact of a 28-day supplementation with rhLF at two doses compared to an active control product formulated with bLF on the development of anti-lactoferrin antibodies and iron homeostasis biomarkers in healthy adults. Study 1 is the first study of its kind to directly assess the alloimmunization/immunogenicity potential of an orally ingested food protein (rhLF) by assessing for the breakdown of tolerance following ingestion of an exogenous human protein. During the method development for the anti-LF antibody assay for Study 1, high variability in anti-LF (human and bovine) antibodies was observed in free-living adults. Therefore, Study 2 was designed to characterize the biological variability of anti-LF antibodies (bovine and human) over a 12-week period in the absence of supplemental lactoferrin in healthy adults.

## 1. MATERIALS AND METHODS

### 2.1 Study Design

Study 1 was a randomized, double-blind, parallel arm, controlled study (NCT06012669) with a screening visit (Day -7), baseline visit (Day 0), and three follow-up vists (Days 28, 56, and 84) **(Figure 1)**. Participants were randomly assigned to one of three 28-day interventions: 3.4 g/d of rhLF as a powder mixed into water (high-dose rhLF), 0.34 g/d of rhLF as a powder mixed into water (low-dose rhLF), or 3.4 g/d of bLF as a powder mixed into water (bLF). Data collection took place between September 2023 and December 2023 in two locations: Health Awareness (Port St. Lucie, FL) and Suncoast Research (Miami, FL). Study 2 was a separate observational study with a screening/baseline visit on Day 0 and three follow-up visits on Days 28, 56, and 84 **(Figure 1)**. No intervention was provided during Study 2. Data collection took place between October 2023 and January 2024 at Excellence Medical and Research (Miami Gardens, FL). The procedures followed were in accordance with the ethical standards on human experimentation, and both studies were approved by the WIRB-Copernicus Group Institutional Review Board (protocol: MB-2305, 8/16/2023; protocol: MB-2310, 9/29/2023).

**Figure 1.**
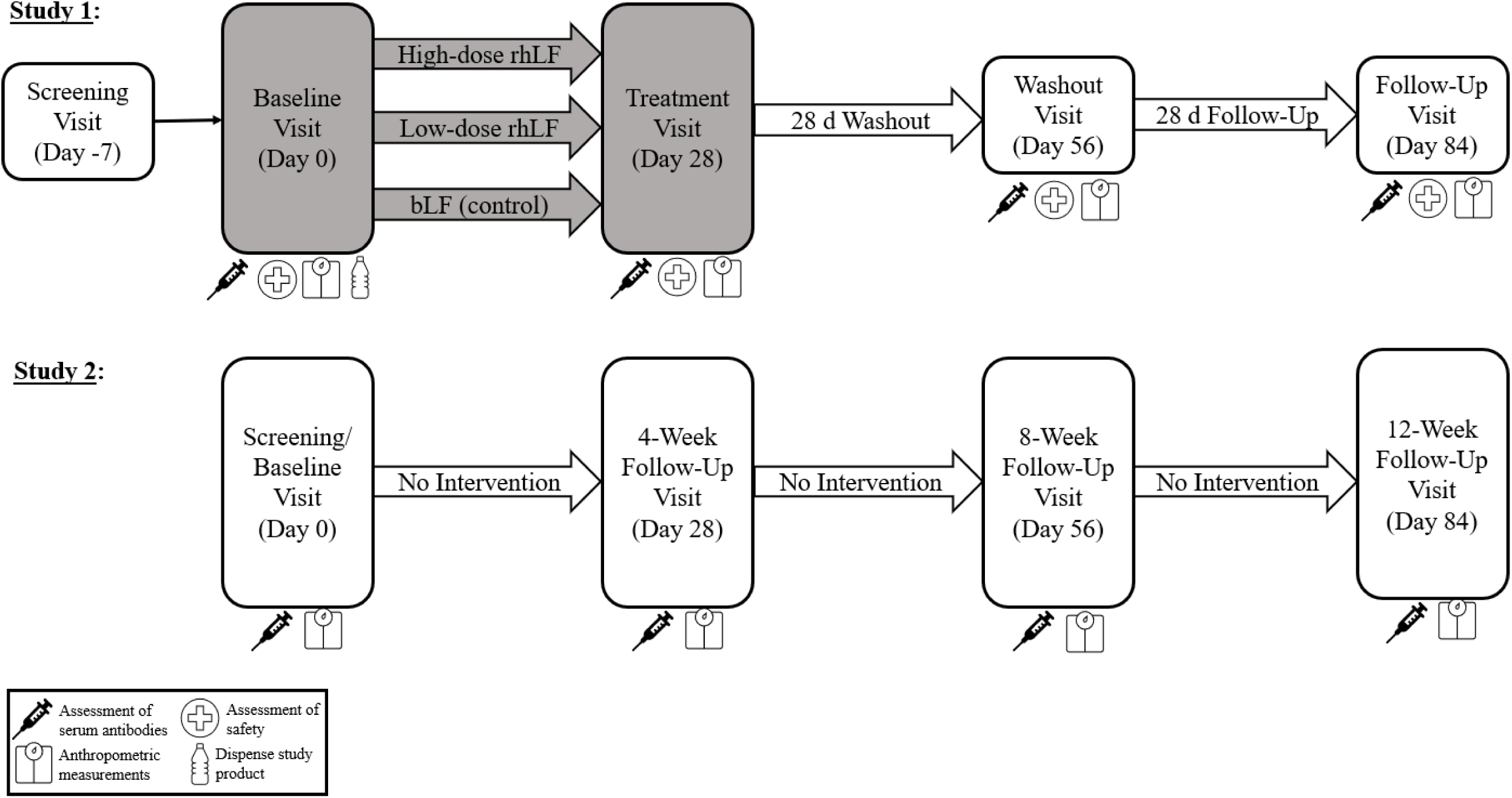
Schematic drawing of the study design for Study 1 and Study 2. Assessments of safety in Study 1 included measurements of iron-related outcomes, hs-CRP, chemistry profile, and complete blood count in serum or plasma, and a urinalysis. Abbreviations: bLF, bovine lactoferrin rhLF, recombinant human lactoferrin.

### 2.2 Participants

Inclusion and exclusion criteria were similar for Study 1 and Study 2. Healthy males and premenopausal females between the ages of 18 and 45 years of age with a body mass index (BMI) of 18.5 to 29.9 kg/m^2^ were recruited. Since the mean BMI in the United States was 29.9 kg/m^2^ in 2017-2018 (26), this BMI range better represents healthy adults without disease. Health status was assessed based on medical history and routine laboratory tests. Participants in each study agreed to not receive any type of vaccination within 7 days of the screening visit through Day 56. The screening visit took place on Day -7 in Study 1 and Day 0 in Study 2. Prior to testing visits, participants abstained from alcohol and vigorous physical activity for at least 24 hours.

Participants were excluded if taking medications or over-the-counter products that could interfere with the absorption and metabolism of lactoferrin or affect study-related outcomes within 14 days of Day 0 in both studies. Examples of these medications include aspirin, anti-inflammatory medications, antacids, histamine-2 receptor antagonists, laxatives, stool softeners, and proton pump inhibitors. Individuals taking herbal and dietary supplements within 14 days of Day 0 were also excluded from each study, except that multivitamin, mineral, and calcium supplements were permitted. Participants were also excluded if presenting with signs/symptoms of an active infection or taking antibiotics within 5 and 7 days prior to any visit, respectively. A washout period was permitted for dietary supplements, infections, and antibiotics. Furthermore, a history of bariatric surgery, current use of any weight loss drug, weight change of 4.5 kg (10 lbs.) within 3 months of the screening visit, extreme dietary habits, and intention to lose or gain body weight were exclusionary.

Individuals with autoimmune disorders and gastrointestinal conditions that could interfere with the absorption of lactoferrin were excluded from both studies. Other exclusion criteria included chronic nicotine use, a laboratory test result of clinical significance, a positive result on a urine drug screen, a clinically significant medical diagnosis, uncontrolled hypertension (systolic blood pressure ≥160 mm Hg and/or diastolic blood pressure ≥100 mm Hg), allergies or sensitivities to any component of the study products or yeast, and a history of cancer within 2 years (except non-melanoma skin cancer or carcinoma in situ of the cervix).

Additionally, participants were excluded if they had been exposed to any non-registered drug product within 30 days of the screening visit or if they had history of drug or alcohol abuse within 12 months of the screening visit. Females were excluded if they were postmenopausal, pregnant, planning to become pregnant during the study period, lactating, or of childbearing potential without a medically approved form of contraception during the study period. Lastly, the investigator excluded participants who had a medical condition that would interfere with the participant’s ability to provide informed consent, comply with the study protocol, confound the interpretation of the results, or create undue risk for the participant. Participants were not permitted to participate in both studies 1 and 2.

### 2.3 Protocol

#### 2.3.1 Screening/Baseline Visits

In Study 1, the screening visit (Day -7) and the baseline visits (Day 0) were conducted separately, while these visits were conducted together on Day 0 in Study 2. For the screening visit in both studies, participants arrived at the clinic following a 9-14 hour overnight fast. After informed consent was provided, medical history during the past 5 years and prior/current medication use was evaluated. Height and body weight were measured with calibrated scales and stadiometers. Vital signs were also measured. BMI was calculated to confirm eligibility (18.5 to 29.9 kg/m^2^ was inclusionary). A fasting venous blood sample was collected for a chemistry profile and complete blood count (CBC) in both studies and for serum anti-LF antibodies in Study 2 only. A urine sample was collected for a pregnancy test (females only), urine drug screen, and a urinalysis. AEs were assessed through open-ended questions and spontaneous reporting. Participants were instructed to avoid alcohol and vigorous physical activity for 24 h prior to all visits. Two 3-day diet records were dispensed to assess dietary intake (2 weekdays and 1 weekend day or holiday) at baseline (prior to any intervention if applicable) and during week 4.

Participants in Study 2 did not receive an intervention and were scheduled to return approximately 28 days later. Converesely, if participants in Study 1 qualified based on measures at the screening visit, participants returned to the clinic 2 to 28 days later for the baseline visit (visit 2, Day 0). The 3-day diet record was collected. Concomitant medications were re-evaluated, inclusion/exclusion criteria were reviewed, and body weight and vital signs were measured. Eligible participants were randomly assigned to receive one of the three treatment groups (high-dose rhLF, low-dose rhLF, or bLF). A fasting blood sample was drawn at t = −0.5 hour for analysis of antibodies, high-sensitivity C-reactive protein (hs-CRP), and iron-related outcomes in serum. Next, participants consumed one serving of their assigned study product in the clinic, which was consumed within 30 minutes. One serving of the high-dose rhLF and low-dose rhLF treatments was equivalent to 1.7 and 0.17 g of rhLF, respectively. One serving of the bLF treatment was equivalent to 1.7 g of bLF. Additional study products based on randomization assignment and a study product compliance log were dispensed. AEs were assessed. Participants were instructed to take two servings of the assigned product every day until the next clinic visit. In addition, participants were instructed to maintain their regular physical activity pattern.

#### 2.3.2 Follow-up Visit on Day 28

In both studies, participants returned to the clinic on Day 28 following a 9-14 hour fast and 24 hours without vigorous physical activity or alcohol consumption. Compliance with study instructions was verified. Concomitant medications were re-evaluated, study continuation requirements were reviewed, and the 3-d diet record and study product compliance log were collected and reviewed. Body weight and vital signs were measured, and AEs were assessed. In Study 1, blood and urine samples were collected for analyses of serum antibodies, safety-related outcomes (iron-related outcomes, hs-CRP chemistry profile, and CBC) in serum or plasma, and a urinalysis. Also, the leftover study products were collected, and compliance was assessed. In Study 2, a blood sample was collected for analysis of serum antibodies only.

#### 2.3.3 Follow-up Visits on Day 56 and 84

In both studies, participants arrived at the clinic on Days 56 and 84 under the same conditions as the previous visit. The visit window for all follow-up visits was ±2 days. Concomitant medications were assessed, continuation requirements were reviewed, AEs were assessed, and body weight and vital signs were measured. In Study 1, blood and urine samples were collected for the same analyses that occurred on Day 28. In Study 2, another blood sample was collected for analysis of serum antibodies only.

### 2.4 Study Products

In Study 1, the participants were randomized into one of the three treatment groups: high-dose rhLF, low-dose rhLF, or bLF (active control). A blinded statistician generated the randomization scheme, which was stratified by sex (self-reported), using a random number generator. Randomization codes were concealed in sequentially numbered, opaque envelopes until the time of intervention assignment. Blinded staff enrolled and assigned participants to interventions. The participant allocation ratio was 1:1:1, with an equal balance of males and females in each group. The high-dose rhLF treatment group consumed 3.4 g/d (1.7 g/serving) of rhLF, and the low-dose rhLF treatment group consumed 0.34 g/d (0.17 g/serving) of rhLF. The bLF treatment group consumed 3.4 g/d (1.7 g/serving) of bLF. One serving of the powder study product was dissolved into 12 to 16 oz. of drinking water directly prior to consumption. Two servings of study products were consumed 8-12 h apart each day, and each dose was consumed within 30 minutes. The study products were manufactured by National Food Lab (Naples, FL). The study products contained maltodextrin, natural flavor, citric acid, fruit juice, vegetable juice, sucralose, and either rhLF or bLF. The nutrient composition of one serving of each study product is provided in **Supplemental Table 1**. Compliance with product consumption was assessed by counting the number of sachets returned to the clinic and reviewing the study product compliance logs.

In Study 2, no study products were provided due to the observational nature of the study.

### 2.5 Laboratory Measurements and Other Procedures

For both studies, the primary outcome variable was the change in anti-lactoferrin antibodies in serum from Day 0 to Day 56, and the exploratory outcome variable was the change in serum anti-lactoferrin antibodies from Day 0 to Days 28 and 84. Change from baseline for serum anti-lactoferrin antibodies was expressed as the ratio of the post-exposure value (Day 28, 56, or 84) to the Day 0 value. For Study 1, the secondary outcome variables included analysis of PBMC with and without ex vivo stimulation with lactoferrin and will be reported elsewhere.

#### 2.5.1 Anti-Lactoferrin Antibodies in Serum

Two assays were developed and evaluated at Immunologix Laboratories (Tampa, FL). Both assays were developed to detect anti-lactoferrin antibodies according to the recommendations in the FDA Guidance for Industry, Immunogenicity Testing of Therapeutic Protein Products — Developing and Validating Assays for Anti-Drug Antibody Detection (22). Although LF is not a drug therapy, the FDA guidance outlines recommendations for assays to reliably detect anti-target protein antibodies that may be associated with clinical events. One assay detected antibodies against bLF, while the other assay detected antibodies against hLF. The level of sensitivity for both assays was at least 100 ng/mL as defined by commercially available anti-bLF or anti-hLF antibodies used for characterization of the assays. Sensivity of at least 100 ng/mL is recommended by the FDA for detection of antibodies that may be associated with clinical events.

#### 2.5.2 Anti-bovine Lactoferrin Antibody Assay

This method is a bridging electrochemiluminescence (ECL) screening assay to detect anti-bLF antibodies in human serum. Due to high prevalence of lactoferrin-specific signals from normal healthy individuals, the increase in signal of post-exposure samples relative to baseline (pre-exposure) signal was used to determine study product-emergent anti-bLF antibodies. Pre-exposure samples were tested on the same plate as post-exposure samples to generate a post/pre ratio (i.e. raw signal of post divided by raw signal of pre). In the assay, biotinylated bLF was added to a streptavidin Gold Mesoscale Discovery (MSD) plate at 1 µg/mL in phosphate-buffered saline (PBS) at ambient temperature with shaking to coat the plate with biotinylated bLF. Following coating, the plate was washed and then blocked with Superblock (Thermo Fisher) at ambient temperature with shaking. Samples and controls were diluted 1:4 in assay diluent (2% SuperBlock in PBS) within a transfer plate. Samples and controls were further diluted 1:5 with 100 mM acetic acid and allowed to incubate at ambient temperature with shaking. Blocked streptavidin plates were then washed to remove any unbound material. After the acid treatment, samples and controls were neutralized with neutralization buffer (150 mM Tris) for a final minimum required dilution (MRD) of 1:30 and immediately plated onto the blocked MSD plate and allowed to incubate at ambient temperature with shaking. After incubation, the plates were washed of any unbound material, and a detector solution containing SULFO-TAG^TM^ conjugated bLF was added at 1 µg/mL in assay diluent and allowed to incubate at ambient temperature with shaking. After incubation, the plate wells were washed, and read buffer (MSD Read Buffer T) was added to the wells of the plate. The raw signal (ECL units) was measured using an MSD Sector Imager. Raw signal from post-exposure samples were divided by pre-exposure raw signal from the same individual, resulting in the post/pre ratio. Increases in post/pre ratios indicate the presence of product-emergent anti-bLF antibodies.

#### 2.5.3 Anti-human Lactoferrin Antibody Assay

A bridging ECL screening assay was also used to detect anti-hLF antibodies in human serum. Due to high prevalence of lactoferrin-specific signals from normal healthy individuals, the increase in signal of post-exposure samples relative to baseline (pre-exposure) signal was used to determine study product-emergent anti-hLF antibodies. Pre-exposure samples were tested on the same plate as post-exposure samples to generate a post/pre ratio (i.e. raw signal of post divided by raw signal of pre). In the assay, biotinylated hLF was added to a streptavidin Gold MSD plate at 1 µg/mL in PBS at ambient temperature with shaking to coat the plate with biotinylated hLF. Following coating, the plate was washed and then blocked with Superblock (Thermo Fisher) at ambient temperature with shaking. Samples and controls were diluted 1:2 in assay diluent (2% SuperBlock in PBS) within a transfer plate. Samples and controls were further diluted 1:5 with 100 mM acetic acid and allowed to incubate at ambient temperature with shaking. Blocked streptavidin plates were then washed to remove any unbound material. After the acid treatment, samples and controls were neutralized with neutralization buffer (150 mM Tris) for a final MRD of 1:15 and immediately plated onto the blocked MSD plate and allowed to incubate at ambient temperature with shaking. After incubation, the plates were washed of any unbound material, and a detector solution containing SULFO-TAG^TM^ conjugated hLF was added at 1 µg/mL in assay diluent and allowed to incubate at ambient temperature with shaking. After incubation, the plate wells were washed, and read buffer (MSD Read Buffer T) was added to the wells of the plate. The raw signal (ECL units) was measured using an MSD Sector Imager. Raw signal from post-exposure samples were divided by pre-exposure raw signal from the same individual, resulting in the post/pre ratio. Increases in post/pre ratios indicate the presence of product-emergent anti-hLF antibodies.

#### 2.5.4 Safety Assessments

For Study 1, chemistry profile, CBC, and urinalysis at Day -7, Day 28, Day 56, and Day 84 and the iron-related outcomes and hs-CRP at Day 0, Day 28, Day 56, and Day 84 were assessed by LabCorp (Hollywood, FL; Tampa, FL). The chemistry profile included analysis of glucose, blood urea nitrogen, creatinine, estimated glomerular filtration rate, sodium, potassium, carbon dioxide, calcium, total protein, albumin, total globulin, bilirubin, aspartate transaminase, and alanine transaminase. For the CBC, blood was collected in lavender top ethylenediaminetetraacetic acid (EDTA) tubes and refrigerated until analysis of white and red blood cell counts, hemoglobin, hematocrit, mean corpuscular volume, mean corpuscular hemoglobin concentration, and a platelet count with reflex differential. For the urinalysis, urine was collected in a urine collection cup before being transferred into a urinalysis tube with preservative and refrigerated until analysis. The urinalysis included analyses for specific gravity, pH, urine color and appearance, white blood cell esterase, protein, glucose, ketones, occult blood, bilirubin, urobilinogen, and nitrite.

For Study 1, blood for measurements of iron, ferritin, iron saturation, unsaturated iron binding capacity (UIBC), total iron binding capacity (TIBC), and hs-CRP was collected in an SST before clotting upright at room temperature for 30 minutes and centrifuging for 10 minutes at 1100-1300 g to separate the serum for analysis. Iron, iron saturation, UIBC, and TIBC were analyzed using colorimetric assays, and ferritin was analyzed using an electrochemiluminescence immunoassay. For soluble transferrin receptor, blood was collected in a lavender top EDTA tube, centrifuged for 10 minutes at 1100-1300 g, then all plasma was transferred into a transfer tube and stored in the refrigerator until analysis via immunochemiluminometric assay.

Also for Study 1, the number of product-emergent AEs, defined as AEs that occurred during or after the first dose of study product, and the number of participants who experienced at least one product-emergent AE were safety outcomes.

For Study 2, eligibility for the study on Day 0 were evaluated by the chemistry profile, CBC, and urinalysis that were conducted by InterLab (Doral, FL).

#### 2.5.6 Urine Drug Screen

An in-clinic urine drug screening test using the iCasette Dx Drug Screen (Alere Inc., Waltham, MA) was completed at each study’s screening visit. The drug screening included assessment of amphetamines, cocaine, marijuana, benzodiazepines, tricyclic antidepressants, barbiturates, ecstasy (MDMA phencyclidine), oxycodone, and propoxyphene.

### 2.6 Statistical Analysis

Statistical analyses were conducted using R Statistical Software (v4.2.2; R Core Team 2022) and followed a pre-specified statistical analysis plan. For both studies, baseline demographics and clinical characteristics were summarized for the overall sample. Continuous variables were presented as mean and SD if normally distributed or as median and interquartile range limits. Categorical variables are presented as the number and the percentage of participants in each category.

#### 2.6.1 Study 1

For Study 1, an evaluable sample of 60 participants (∼50% for each sex, where sex is defined as sex assigned at birth) is expected to provide at least 80% power to detect a difference in change from baseline of 1.5 standard deviations for each sex subgroup, compared with the corresponding active control subgroup, for key outcomes without adjustment for the number of variables tested. A sample of 66 participants was randomized to allow for ∼10% attrition and/or non-compliance. The intent-to-treat (ITT) population included all participants who were randomized to a study product. The ITT analysis was the primary analysis. The safety population included all participants in the ITT population who consumed any amount of study product. The safety population was the analytical population for all safety outcomes. The per protocol population included all participants in the ITT population who were compliant with the assigned study product, and for whom no clinically important protocol deviations occurred during the 8-week treatment period. Participants who consumed 80-120% (inclusive) of the scheduled doses were deemed compliant with the study product. All decisions regarding inclusion in the per protocol datasets were made and documented prior to database lock and unblinding. The per protocol analysis was the secondary analysis.

Multiple imputation (MI) was used in the ITT analysis to impute missing data through day 56. The MI procedure used the Markov chain Monte Carlo (MCMC) method with a single chain to create at least 5 imputations or more, in order to obtain a stable result. Each iteration was analyzed, and the final result reported is the summarized values. When values were reported as below the limit of quantitation, they were set to the midpoint between zero and the quantitation limit and included in the summaries and analyses.

The primary outcome of Study 1 was evaluated using an analysis of covariance (ANCOVA) model that included the change from baseline (day 0) to day 56 in serum anti-lactoferrin antibodies as the dependent variable, sex and study product as main effects, a sex-by-product interaction, and baseline as a covariate. A sex-by-product interaction was not observed (p ≥ 0.10) and the interaction term was dropped form the model. Differences between each rhLF group and the bLF group were assessed within the overall sample using a pre-determined hierarchical approach. Comparison of the high-dose rhLF group to the bLF group was tested at α = 0.05, 2 sided, significance level. Comparison of the high-dose rhLF group to the low-dose rhLF group was tested at α = 0.0253, 2-sided, significance level. Comparison of the low-dose rhLF group to the bLF group was tested at α = 0.0167, 2-sided, significance level. Assumptions of normality of residuals and homogeneity of variance were tested by Shapiro Wilk test and Levene’s test, respectively, using a significance level of 0.01 for both. Due to a violation of model assumptions, a natural log (ln)-transformation was applied to the dependent variable. Normality of the residuals was not achieved with the ln transformation and a ranked analysis was also performed. Since there were no material differences between the two models, least squares geometric means (LS GM) and their 95% CIs were reported from the ln-transformed model and p-values were reported from the ranked model.

The exploratory outcomes of Study 1 were evaluated using separate ANCOVA models for days 28, 56, and 84. Initial ANCOVA models were similar to those described for the primary outcome. Significant sex-by-product interactions were not observed for any of the explorary outcomes (p ≥ 0.10) and the interaction term was dropped from the models. Differences between each rhLF group and the bLF group were assessed within the overall sample using an α = 0.05, two-sided, level of significance. There was no adjustment for the testing of multiple outcomes or multiple comparisons to reduce the risk of making a type II error.

Post hoc exploratory outcomes included changes in the pre-specified primary and exploratory outcomes from baseline (day 0) to days 28, 56, and 84 (when appropriate) within each sex-product subgroup. Changes from baseline within each sex-product subgroup and differences between product groups within each sex subgroup were assessed using descriptive statistics, including 95% CIs for the mean.

The frequency of product-emergent AEs and participants who experienced at least one product-emergent AE were summarized by relationship to the study product (not related, unlikely, possibly, probably, or definitely related to the study product) and severity (mild, moderate, or severe). An AE’s relationship to the study product and severity were determined by the investigator. Multiple occurrences of the same event were counted only once per participant. Differences between product groups in the frequency of product-emergent AEs and participants who experienced at least one product-emergent AE were assessed using chi-square tests or Fisher’s exact test as appropriate. A similar analysis was planned for product-emergent AEs that were related to a study product, defined as those possibly, probably, or definitely related to the study product. However, the analysis was not performed due to the low frequency of product-emergent AEs that were deemed related to a study product. For analytes from the chemistry panel, hematology, and urinalysis and the iron-related outcomes, comparisons between each rhLF group and the bLF group were performed using descriptive statistics, including 95% CIs.

#### 2.6.2 Study 2

For Study 2, twenty-four healthy adults were enrolled. The evaluable population included all participants who were enrolled in the study and provided data for at least one post-enrollment visit. The per protocol population included all participants who are included in the evaluable population and for whom no clinically important protocol deviations occurred during the 8-week follow-up period. The evaluable analysis was the primary analysis, and the per protocol analysis was the secondary analysis. Only observed data was included in the analyses. The change and variability in the primary and secondary outcomes were assessed using descriptive statistics. The LS GMs and 95% CIs for the serum anti-LF antibody post/pre ratio at each time point was obtained from separate mixed linear models that included assay as a fixed effect, baseline (untransformed) as a covariate, and participant as a random effect.

## 2. RESULTS

In Study 1, eighty-four individuals were screened, and sixty-six were randomly assigned to a study product (n = 23, bLF; n = 21, low-dose rhLF; n = 22, high-dose rhLF). Nine participants withdrew consent (n = 3 per group), one participant in the bLF group was lost to follow-up, and one participant in the high-dose rhLF group dropped out due to a serious AE that was unrelated to the study product **(Figure 2)**. Four additional participants were excluded from the per protocol analysis due to excluded medication use during the study period (n = 1 bLF; n = 1 high-dose rhLF) and compliance <80% (n = 1 low-dose rhLF; n = 1 high-dose rhLF).

**Figure 2.**
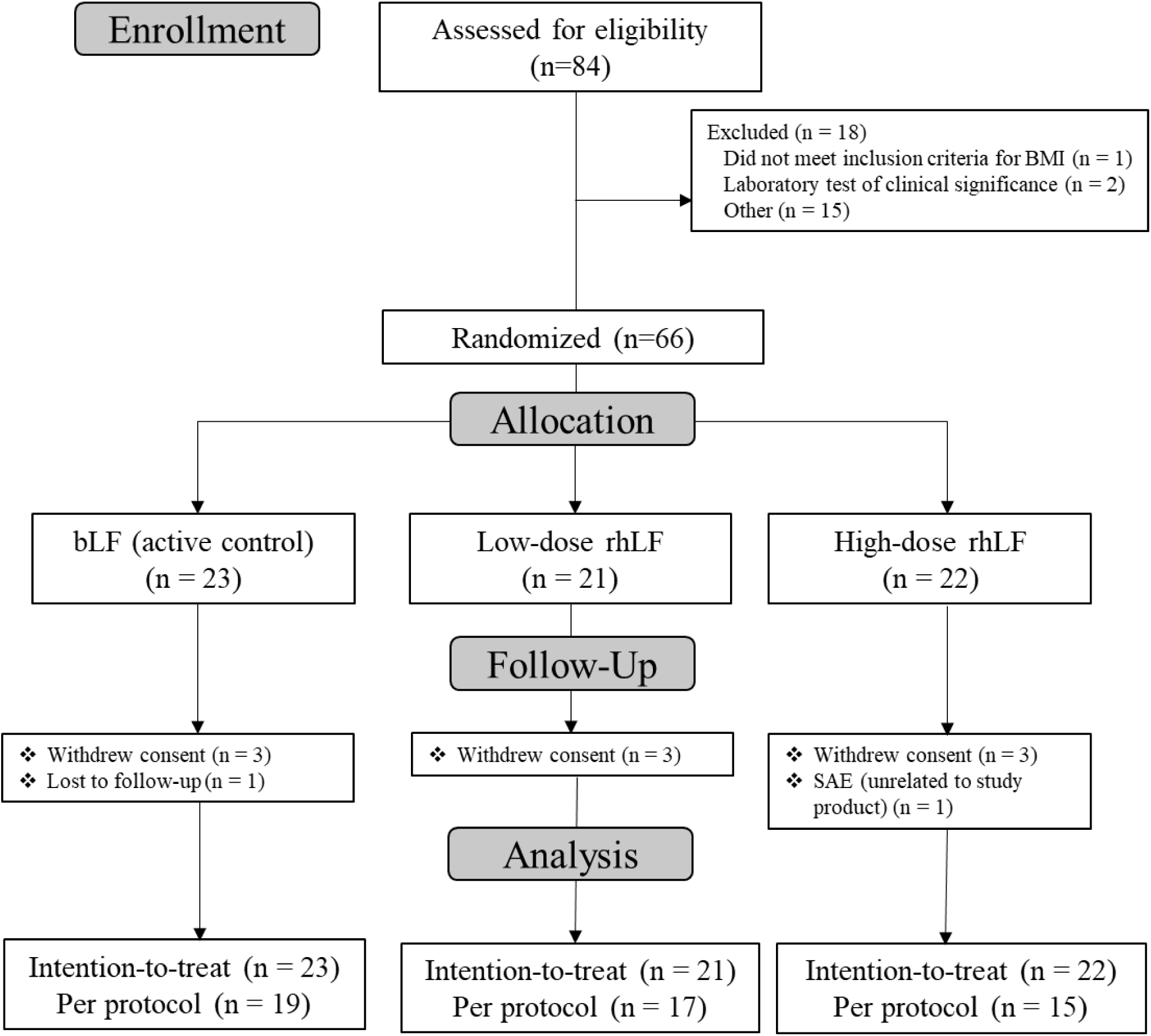
Consolidated Standards of Reporting Trials (CONSORT) flow diagram of selection of participants in Study 1.

Therefore, fifty-one participants were included in the per protocol analysis (n = 19 bLF; n = 17 low-dose rhLF; n = 15 high-dose rhLF) and all randomized participants were included in the full analysis set using the ITT principle. The demographic and baseline characteristics of the 66 randomized participants are presented in **Table 1**. The mean age and BMI of all participants were 36.9 ± 7.80 years and 26.5 ± 2.80 kg/m^2^, respectively. Eighty-two percent of the participants identified as Hispanic/Latino. In the ITT population, mean (± SE) compliance was 97.7 ± 1.5%, 97.6 ± 2.2%, and 93.7 ± 3.8% in the bLF (n = 21), low-dose rhLF (n = 18), and high-dose rhLF (n = 20), respectively, which included data from participants that completed the 28-d intervention. In the per protocol population, compliance was 97.8 ± 1.6%, 99.8 ± 0.4%, and 98.4 ± 1.8% in the bLF, low-dose rhLF, and high-dose rhLF, respectively.

**Table 1.** Characteristics of participants in Study 1 at baseline in the intention-to-treat population.

| Characteristic | Value |  |  |  |  |  |
| --- | --- | --- | --- | --- | --- | --- |
|  | Bovine Lactoferrin<br>(Active Control) |  | Low-Dose Recombinant<br>Human Lactoferrin |  | High-Dose Recombinant<br>Human Lactoferrin |  |
|  | Female (n = 11) | Male (n = 12) | Female (n = 10) | Male (n = 11) | Female (n = 11) | Male (n = 11) |
| Race, no. (%) |  |  |  |  |  |  |
| White | 11 (100) | 8 (66.7) | 7 (70.0) | 10 (90.9) | 10 (90.9) | 9 (81.8) |
| Black/African American | 0 (0) | 3 (25.0) | 2 (20.0) | 1 (9.09) | 1 (9.09) | 2 (18.2) |
| American Indian/Alaskan Native | 0 (0) | 0 (0) | 1 (10.0) | 0 (0) | 0 (0) | 0 (0) |
| Declined to Specify | 0 (0) | 1 (8.33) | 0 (0) | 0 (0) | 0 (0) | 0 (0) |
| Ethnicity, no. (%) |  |  |  |  |  |  |
| Hispanic/Latino | 8 (72.7) | 9 (75.0) | 9 (90.0) | 10 (90.9) | 9 (81.8) | 9 (81.8) |
| Non-Hispanic/Latino | 3 (27.3) | 3 (25.0) | 1 (10.0) | 1 (9.09) | 2 (18.2) | 2 (18.2) |
| Nicotine Use, no. (%) |  |  |  |  |  |  |
| Never Used | 11 (100) | 11 (91.7) | 10 (100) | 11 (100) | 11 (100) | 11 (100) |
| Current User | 0 (0) | 1 (8.33) | 0 (0) | 0 (0) | 0 (0) | 0 (0) |
| Alcohol Use, no. (%) |  |  |  |  |  |  |
| None | 7 (63.6) | 7 (58.3) | 4 (40.0) | 7 (63.6) | 5 (45.5) | 5 (45.5) |
| Occasional | 4 (36.4) | 4 (33.3) | 6 (60.0) | 4 (36.4) | 5 (45.5) | 6 (54.6) |
| Weekly | 0 (0) | 1 (8.33) | 0 (0) | 0 (0) | 1 (9.09) | 0 (0) |
| Age, mean (SD), y | 41.1 (10.4) | 33.5 (7.08) | 39.2 (5.42) | 33.5 (7.37) | 36.5 (8.61) | 38.1 (4.97) |
| SBP, mean (SD), mm Hg | 113 (15.1) | 121 (8.22) | 115 (9.25) | 118 (9.36) | 110 (11.6) | 120 (9.51) |
| DBP, mean (SD), mm Hg | 73.1 (12.2) | 75.8 (5.55) | 76.2 (6.80) | 74.7 (6.15) | 71.9 (8.80) | 76.7 (9.17) |
| BMI, mean (SD), kg/m<sup>2</sup> | 26.5 (2.71) | 26.9 (2.39) | 25.9 (2.36) | 25.9 (3.68) | 25.2 (3.26) | 28.3 (1.39) |

BMI, body mass index; DBP, diastolic blood pressure; SBP, systolic blood pressure.

In Study 2, twenty-eight participants were screened, and twenty-four were enrolled. The demographic and baseline characteristics of the participants are presented in **Table 2**. The participant characteristics in Study 2 were very similar to Study 1. For example, the mean age and BMI were 32.8 ± 6.39 years and 26.2 ± 1.98 kg/m^2^, respectively. All twenty-four participants identified as Hispanic/Latino. There were no dropouts or adverse events during the study period.

**Table 2.** Characteristics of study participants at baseline in Study 2.

| **Characteristic** | **Female (n = 12)** | **Male (n = 12)** |
| --- | --- | --- |
| Race, no. (%) |  |  |
| White | 11 (91.7) | 11 (91.7) |
| Black/African American | 1 (8.33) | 1 (8.33) |
| Ethnicity, no. (%) |  |  |
| Hispanic/Latino | 12 (100) | 12 (100) |
| Non-Hispanic/Latino | 0 (0) | 0 (0) |
| Nicotine Use, no. (%) |  |  |
| Never Used | 12 (100) | 11 (91.7) |
| Ex-Nicotine User | 0 (0) | 1 (8.33) |
| Alcohol Use, no. (%) |  |  |
| None | 12 (100) | 1 (8.33) |
| Occasional | 0 (0) | 11 (91.7) |
| Weekly | 0 (0) | 0 (0) |
| Age, mean (SD), y | 32.2 (5.06) | 33.4 (7.68) |
| SBP, mean (SD), mm Hg | 117 (6.16) | 118 (4.51) |
| DBP, mean (SD), mm Hg | 74.2 (7.70) | 71.9 (5.20) |
| BMI, mean (SD), kg/m<sup>2</sup> | 26.3 (1.74) | 26.1 (2.26) |

BMI, body mass index; DBP, diastolic blood pressure; SBP, systolic blood pressure.

### 3.1 Serum Antibodies

The changes from baseline in serum anti-LF antibodies at Day 28, Day 56, and Day 84 for the ITT population in Study 1 and the evaluable population in Study 2 are reported in **Tables 3-4**, **Figure 3**, and **Supplemental Figure 1**. The change from baseline is presented as the post/pre ratio. The post/pre ratio was calculated by dividing the post-exposure sample raw signal by the pre-exposure raw signal. Because the assay signal reflects the presence of anti-LF antibodies, post/pre ratios above 1 indicate an increased presence of anti-LF antibodies. For the primary outcome in Study 1, the increase in serum anti-bLF antibodies in the bLF group was greater than the changes in serum anti-hLF in the low-dose rhLF (P < 0.001) and high-dose rhLF groups (P < 0.001) **(Table 3** and **Figure 3A)**. There were no differences between the two rhLF groups (P = 0.231). The same significant differences were observed in all participants at Days 28 and 84, and when females and males were analyzed separately at Days 28, 56, and 84 **(Supplemental Table 2)**. The changes in serum anti-lactoferrin antibodies in the per protocol population are reported in **Supplemental Table 3**. The findings from the per protocol analysis were not materially different from those in the ITT analysis. In Study 2, median anti-bLF antibodies were 1320 ECL units, and median anti-hLF antibodies were 685 ECL **(Table 4** and **Figure 3B)**. These values did not materially change throughout the course of the study period.

**Figure 3.**
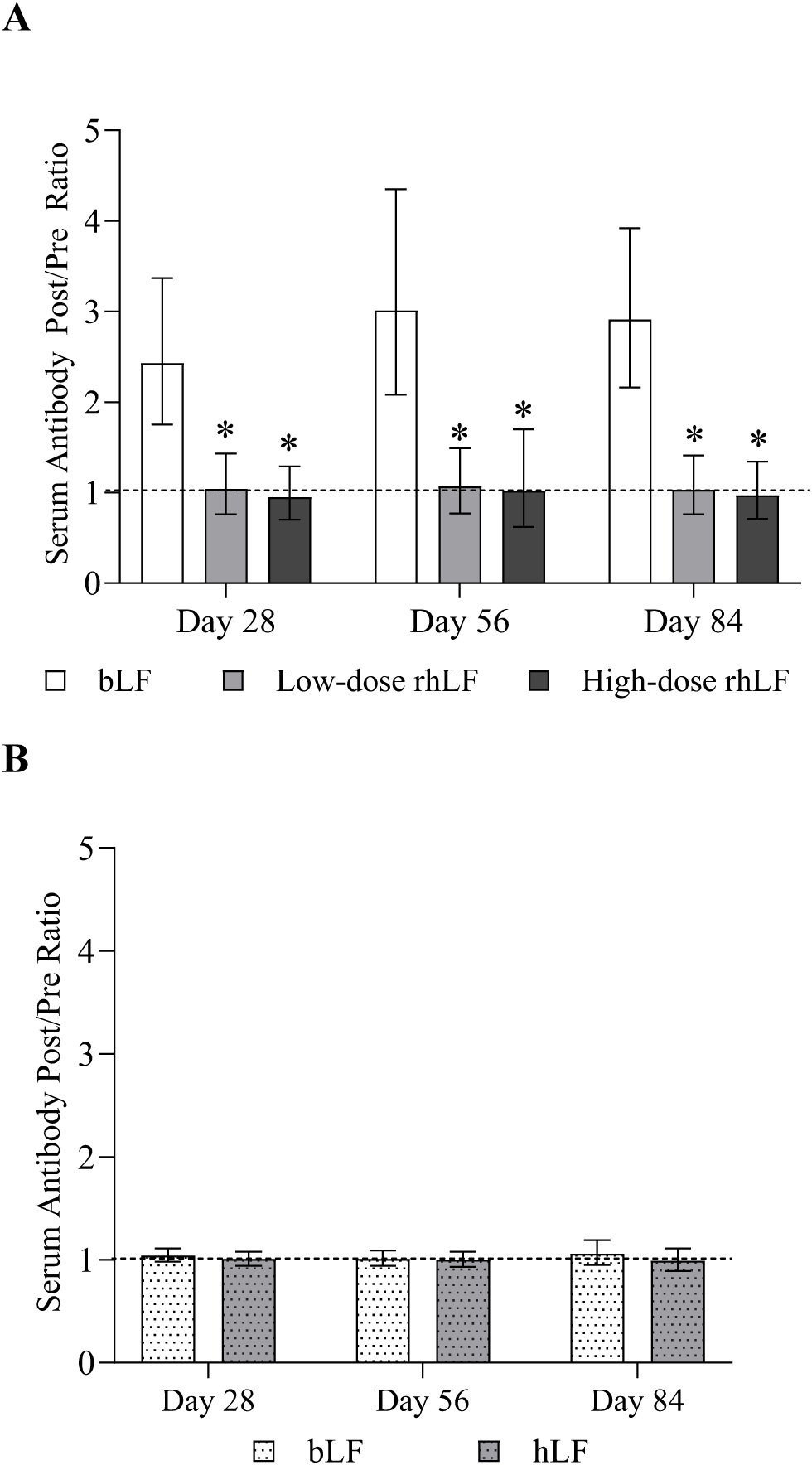
Least squares geometric means and 95% confidence intervals for the change from baseline in serum anti-lactoferrin antibody (expressed as the post/pre ratio) at Days 28, 56, and 84 in A) the bLF, low-dose rhLF, and high-dose rhLF groups in the intention-to-treat population of Study 1 and B) all participants in Study 2. The horizontal dashed line represents a post/pre ratio of 1, which is equivalent to no change from baseline. * Indicates that the change in serum anti-LF antibodies from baseline to the respective day was significantly different from the bLF group (P < 0.01) in Study 1. Abbreviations: bLF, bovine lactoferrin; hLF, human lactoferrin; rhLF, human recombinant lactoferrin.

**Table 3.** Serum anti-lactoferrin antibodies at baseline (Day 0), Day 28, Day 56, and Day 84 in the intention-to-treat population in Study 1^1^.

| **Anti-LF Antibodies in Serum<sup>2</sup>** | **bLF**<br>**(n = 23)** | **Low-Dose rhLF**<br>**(n = 21)** | **High-Dose rhLF**<br>**(n = 22)** | **P-Values** |  |  |
| --- | --- | --- | --- | --- | --- | --- |
|  |  |  |  | **bLF vs.**<br>**Low-Dose**<br>**rhLF** | **bLF vs.**<br>**High-Dose**<br>**rhLF** | **Low-Dose**<br>**vs. High-**<br>**Dose rhLF** |
| Day 0 Median (IQRL) | 594 (414, 852) | 344 (110, 1003) | 366 (197, 1026) |  |  |  |
| Day 28 LS GM (95% CI) for the Post/Pre Ratio | 2.43 (1.75, 3.37) | 1.04 (0.76, 1.43) | 0.95 (0.70, 1.29) | **<0.001** | **<0.001** | 0.568 |
| Day 56 LS GM (95% CI) for the Post/Pre Ratio | 3.01 (2.08, 4.35) | 1.07 (0.77, 1.49) | 1.02 (0.62, 1.70) | **<0.001** | **<0.001** | 0.231 |
| Day 84 LS GM (95% CI) for the Post/Pre Ratio | 2.91 (2.16, 3.92) | 1.03 (0.76, 1.41) | 0.97 (0.71, 1.34) | **<0.001** | **<0.001** | 0.289 |

<sup>1</sup>Missing data at Days 28 and 56 were estimated using multiple imputation. Therefore, sample sizes were lower for Day 84 (n = 20 bLF; n = 18 low-dose rhLF; n = 17 high-dose rhLF). LS GM (95% CI) were obtained from analysis of covariance models that included the natural ln-transformed changes from baseline (expressed as the post/pre ratio) as the dependent variable, product and sex as fixed effects, and the untransformed baseline as a covariate. Non-normality of model residuals persisted with the ln transformations, so a ranked model was also run. There were no material differences between the two models, so nominal p-values are reported from the ranked model, which were not adjusted for multiple comparisons or the testing of multiple study endpoints. For the primary outcome (serum anti-LF antibodies at Day 56), differences between the product groups were tested using a hierarchical approach. Comparison of the high-dose rhLF group to the active control was tested at the $\alpha = 0.05$ , 2-sided, significance level. Comparison of the high-dose to the low-dose rhLF was tested at $\alpha = 0.0253$ , 2-sided, significance level. Comparison of the low-dose recombinant group to the active control was tested at $\alpha = 0.0167$ , 2-sided, significance level. Exploratory outcomes (serum anti-LF antibodies at Days 0, 28, and 84) were tested at $\alpha = 0.05$ , 2-sided, significance level to minimize the risk of a type II error.
<sup>2</sup>Serum anti-bLF antibodies were measured in the bLF group, and serum anti-human LF antibodies were measured in the low- and high-dose rhLF. Units for antibodies in serum are electrochemiluminescent (ECL) units.
Abbreviations: bLF, bovine lactoferrin; IQRL, interquartile range limits; ln, natural log; LS GM, least squares geometric mean; rhLF, recombinant human lactoferrin.

**Table 4.**
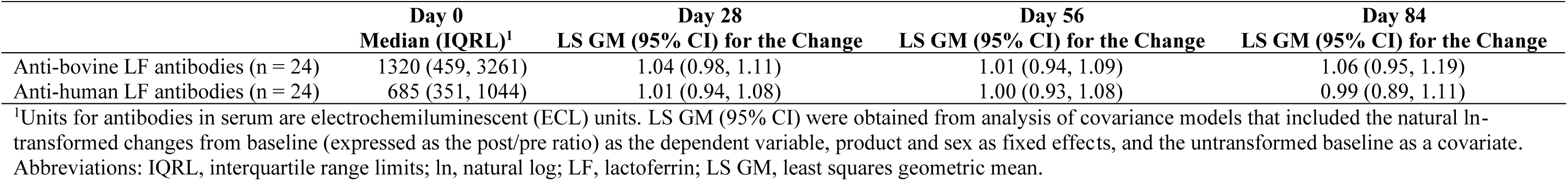
Serum anti-lactoferrin antibodies at baseline (Day 0), Day 28, Day 56, and Day 84 in the evaluable population in Study 2.

Figures 4 and **5** illustrate the post/pre ratio for serum anti-LF antibodies on Day 0 and Day 56 in each individual participant by treatment and sex in Study 1 and Study 2, respectively. The horizontal dashed gray line represents a post/pre ratio of 2, a conservative threshold above which the presence of study product-emergent antibodies is unlikely due to random variability. In Study 1, there was minimal variation between participants in the two rhLF groups, and all ratios were below 2 at Day 56 **(Figure 4B and 4C)**. However, there were twelve participants in the bLF group that had a post/pre ratio greater than or equal to 2 on Day 56, demonstrating that the increase in serum anti-bLF antibodies was shown in more than half of the participants **(Figure 4A)**. More specifically, 7 females in the bLF group had post/pre ratios above 2 on Day 56, and 5 male participants had values above 2. In Study 2, all post/pre ratios were below 2 at Day 56, indicating no material change in any participant **(Figure 5)**.

**Figure 4.**
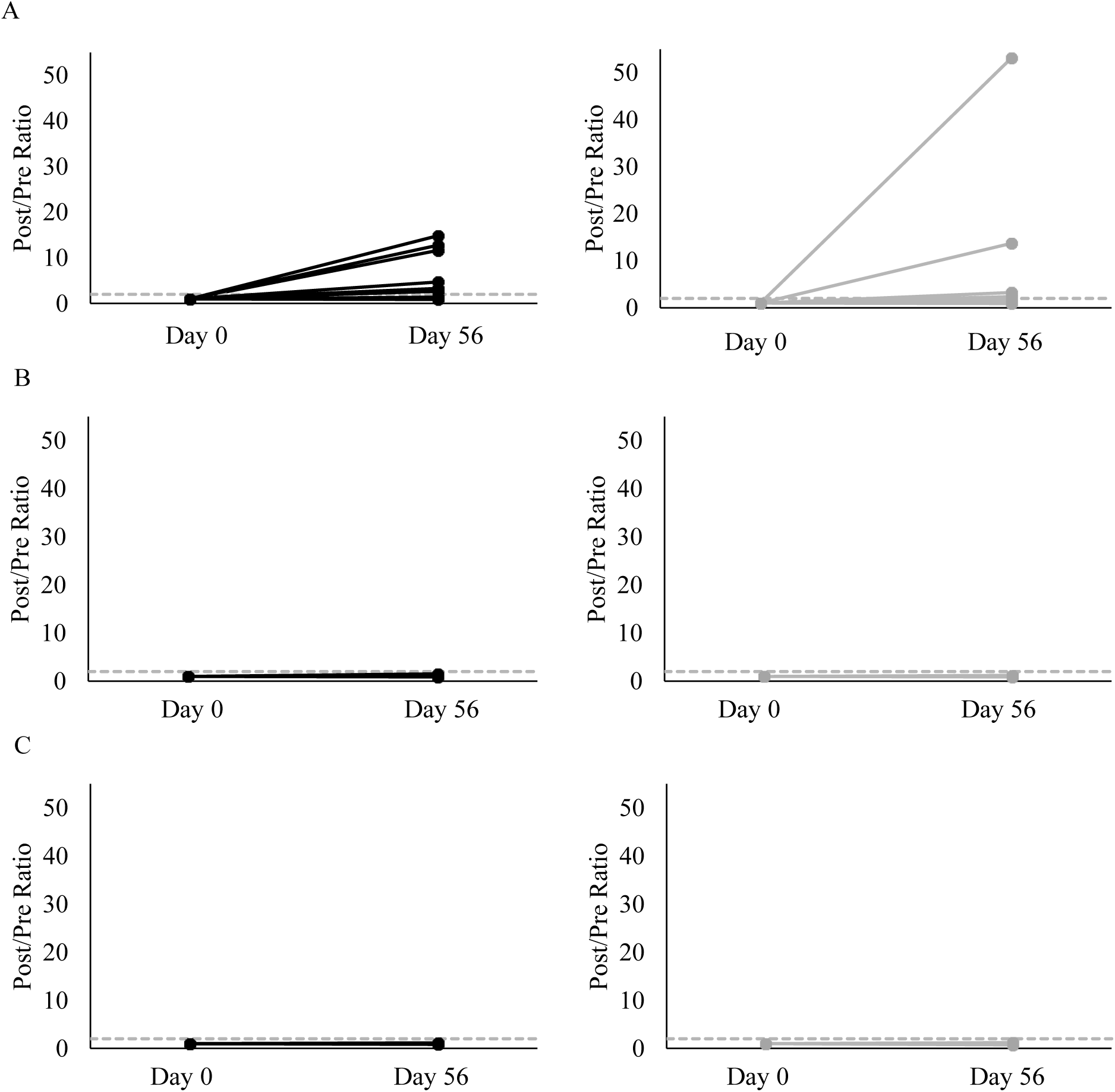
Changes in the post/pre ratios for serum anti-lactoferrin antibodies in females (left panel; solid black lines) and males (right panel; gray lines) at Day 0 and Day 56 in the A) bLF, B) low-dose rhLF, and C) high-dose rhLF groups. The horizontal dashed gray line represents a post/pre ratio of 2, a conservative threshold above which the presence of study product-emergent antibodies is unlikely due to random variability.

**Figure 5.**
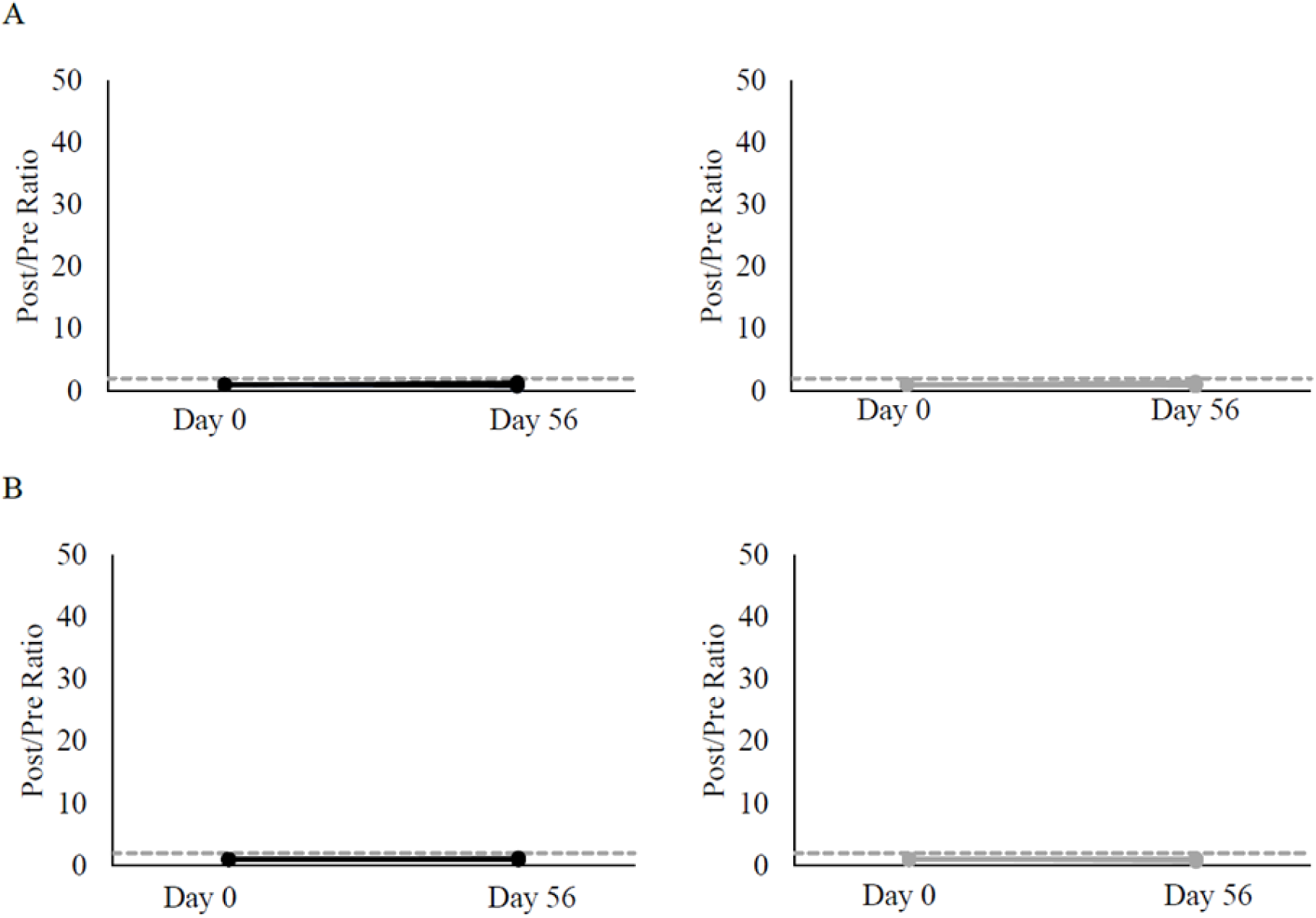
Changes in the post/pre ratios for serum A) anti-bovine lactoferrin antibodies and B) anti-human lactoferrin antibodies at Day 0 and Day 56 in females (left panel; black lines) and males (right panel; gray lines) in Study 2. The horizontal dash gray line represents a post/pre ratio of 2.

### 3.2 Safety Outcomes

Safety outcome analyses for hs-CRP and iron-related measures for Study 1 are reported in the safety population in **Table 5** for all time points. There were no material differences between any of the treatment groups at any time point for hs-CRP, iron, soluble transferrin receptor, ferritin, total iron binding capacity, unsaturated iron binding capacity, and iron saturation. Descriptive statistics for select components of the comprehensive metabolic panel, complete blood count measures and urinalysis are provided in **Supplemental Tables 4-6**. There were no changes within any group for these safety outcomes in the safety population at Day 0, Day 28, Day 56, and Day 84.

**Table 5.**
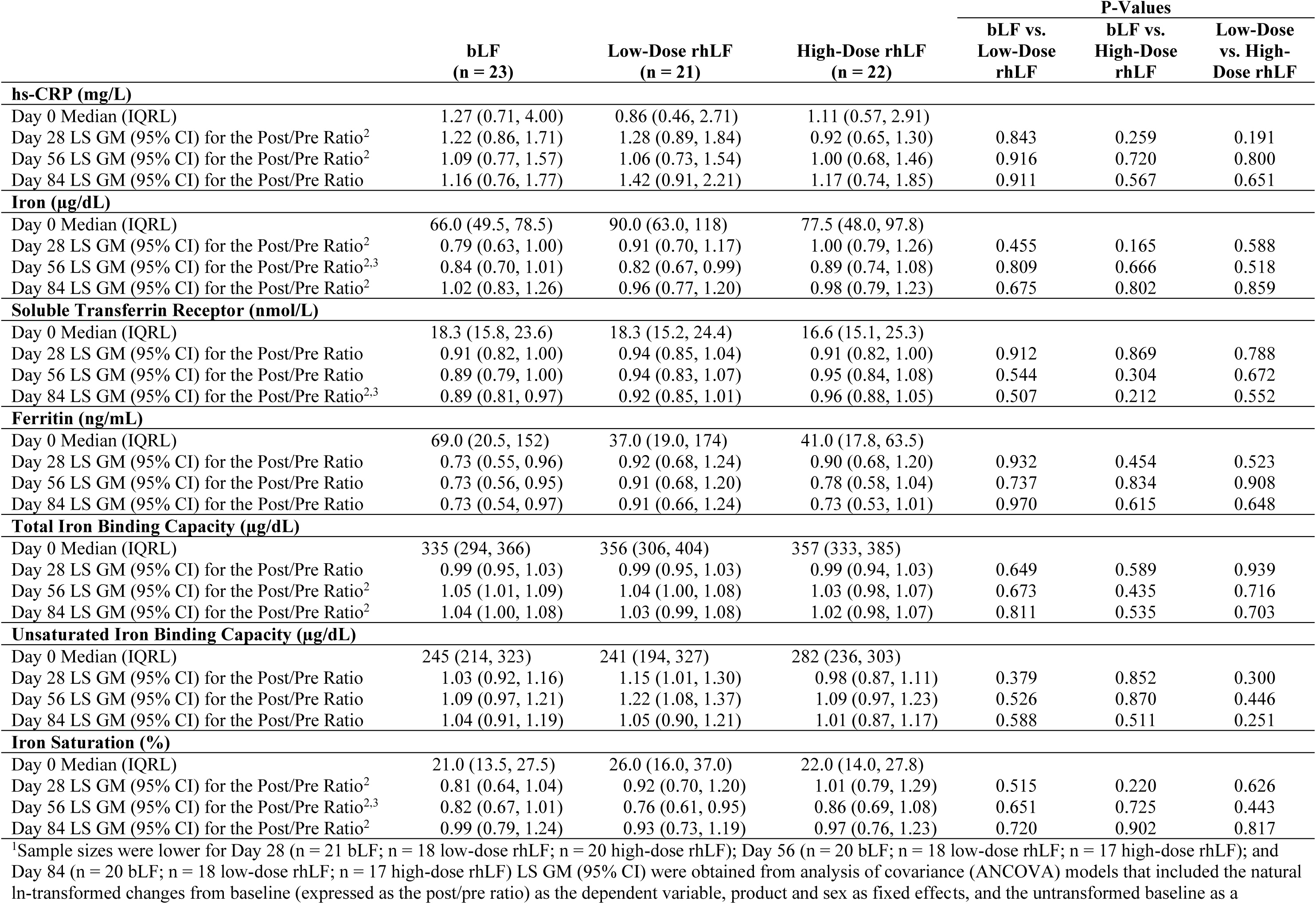
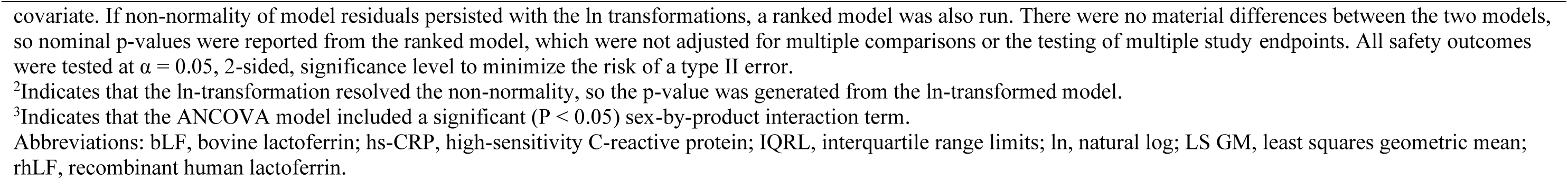
Serum hs-CRP and iron-related outcomes at baseline (Day 0), Day 28, Day 56, and Day 84 in the safety population in Study 1^1^.

|  | bLF<br>(n = 23) | Low-Dose rhLF<br>(n = 21) | High-Dose rhLF<br>(n = 22) | P-Values |  |  |
| --- | --- | --- | --- | --- | --- | --- |
|  |  |  |  | bLF vs.<br>Low-Dose<br>rhLF | bLF vs.<br>High-Dose<br>rhLF | Low-Dose<br>vs. High-Dose rhLF |
| hs-CRP (mg/L) |  |  |  |  |  |  |
| Day 0 Median (IQRL) | 1.27 (0.71, 4.00) | 0.86 (0.46, 2.71) | 1.11 (0.57, 2.91) |  |  |  |
| Day 28 LS GM (95% CI) for the Post/Pre Ratio<sup>2</sup> | 1.22 (0.86, 1.71) | 1.28 (0.89, 1.84) | 0.92 (0.65, 1.30) | 0.843 | 0.259 | 0.191 |
| Day 56 LS GM (95% CI) for the Post/Pre Ratio<sup>2</sup> | 1.09 (0.77, 1.57) | 1.06 (0.73, 1.54) | 1.00 (0.68, 1.46) | 0.916 | 0.720 | 0.800 |
| Day 84 LS GM (95% CI) for the Post/Pre Ratio | 1.16 (0.76, 1.77) | 1.42 (0.91, 2.21) | 1.17 (0.74, 1.85) | 0.911 | 0.567 | 0.651 |
| Iron (µg/dL) |  |  |  |  |  |  |
| Day 0 Median (IQRL) | 66.0 (49.5, 78.5) | 90.0 (63.0, 118) | 77.5 (48.0, 97.8) |  |  |  |
| Day 28 LS GM (95% CI) for the Post/Pre Ratio<sup>2</sup> | 0.79 (0.63, 1.00) | 0.91 (0.70, 1.17) | 1.00 (0.79, 1.26) | 0.455 | 0.165 | 0.588 |
| Day 56 LS GM (95% CI) for the Post/Pre Ratio<sup>2,3</sup> | 0.84 (0.70, 1.01) | 0.82 (0.67, 0.99) | 0.89 (0.74, 1.08) | 0.809 | 0.666 | 0.518 |
| Day 84 LS GM (95% CI) for the Post/Pre Ratio<sup>2</sup> | 1.02 (0.83, 1.26) | 0.96 (0.77, 1.20) | 0.98 (0.79, 1.23) | 0.675 | 0.802 | 0.859 |
| Soluble Transferrin Receptor (nmol/L) |  |  |  |  |  |  |
| Day 0 Median (IQRL) | 18.3 (15.8, 23.6) | 18.3 (15.2, 24.4) | 16.6 (15.1, 25.3) |  |  |  |
| Day 28 LS GM (95% CI) for the Post/Pre Ratio | 0.91 (0.82, 1.00) | 0.94 (0.85, 1.04) | 0.91 (0.82, 1.00) | 0.912 | 0.869 | 0.788 |
| Day 56 LS GM (95% CI) for the Post/Pre Ratio | 0.89 (0.79, 1.00) | 0.94 (0.83, 1.07) | 0.95 (0.84, 1.08) | 0.544 | 0.304 | 0.672 |
| Day 84 LS GM (95% CI) for the Post/Pre Ratio<sup>2,3</sup> | 0.89 (0.81, 0.97) | 0.92 (0.85, 1.01) | 0.96 (0.88, 1.05) | 0.507 | 0.212 | 0.552 |
| Ferritin (ng/mL) |  |  |  |  |  |  |
| Day 0 Median (IQRL) | 69.0 (20.5, 152) | 37.0 (19.0, 174) | 41.0 (17.8, 63.5) |  |  |  |
| Day 28 LS GM (95% CI) for the Post/Pre Ratio | 0.73 (0.55, 0.96) | 0.92 (0.68, 1.24) | 0.90 (0.68, 1.20) | 0.932 | 0.454 | 0.523 |
| Day 56 LS GM (95% CI) for the Post/Pre Ratio | 0.73 (0.56, 0.95) | 0.91 (0.68, 1.20) | 0.78 (0.58, 1.04) | 0.737 | 0.834 | 0.908 |
| Day 84 LS GM (95% CI) for the Post/Pre Ratio | 0.73 (0.54, 0.97) | 0.91 (0.66, 1.24) | 0.73 (0.53, 1.01) | 0.970 | 0.615 | 0.648 |
| Total Iron Binding Capacity (µg/dL) |  |  |  |  |  |  |
| Day 0 Median (IQRL) | 335 (294, 366) | 356 (306, 404) | 357 (333, 385) |  |  |  |
| Day 28 LS GM (95% CI) for the Post/Pre Ratio | 0.99 (0.95, 1.03) | 0.99 (0.95, 1.03) | 0.99 (0.94, 1.03) | 0.649 | 0.589 | 0.939 |
| Day 56 LS GM (95% CI) for the Post/Pre Ratio<sup>2</sup> | 1.05 (1.01, 1.09) | 1.04 (1.00, 1.08) | 1.03 (0.98, 1.07) | 0.673 | 0.435 | 0.716 |
| Day 84 LS GM (95% CI) for the Post/Pre Ratio<sup>2</sup> | 1.04 (1.00, 1.08) | 1.03 (0.99, 1.08) | 1.02 (0.98, 1.07) | 0.811 | 0.535 | 0.703 |
| Unsaturated Iron Binding Capacity (µg/dL) |  |  |  |  |  |  |
| Day 0 Median (IQRL) | 245 (214, 323) | 241 (194, 327) | 282 (236, 303) |  |  |  |
| Day 28 LS GM (95% CI) for the Post/Pre Ratio | 1.03 (0.92, 1.16) | 1.15 (1.01, 1.30) | 0.98 (0.87, 1.11) | 0.379 | 0.852 | 0.300 |
| Day 56 LS GM (95% CI) for the Post/Pre Ratio | 1.09 (0.97, 1.21) | 1.22 (1.08, 1.37) | 1.09 (0.97, 1.23) | 0.526 | 0.870 | 0.446 |
| Day 84 LS GM (95% CI) for the Post/Pre Ratio | 1.04 (0.91, 1.19) | 1.05 (0.90, 1.21) | 1.01 (0.87, 1.17) | 0.588 | 0.511 | 0.251 |
| Iron Saturation (%) |  |  |  |  |  |  |
| Day 0 Median (IQRL) | 21.0 (13.5, 27.5) | 26.0 (16.0, 37.0) | 22.0 (14.0, 27.8) |  |  |  |
| Day 28 LS GM (95% CI) for the Post/Pre Ratio<sup>2</sup> | 0.81 (0.64, 1.04) | 0.92 (0.70, 1.20) | 1.01 (0.79, 1.29) | 0.515 | 0.220 | 0.626 |
| Day 56 LS GM (95% CI) for the Post/Pre Ratio<sup>2,3</sup> | 0.82 (0.67, 1.01) | 0.76 (0.61, 0.95) | 0.86 (0.69, 1.08) | 0.651 | 0.725 | 0.443 |
| Day 84 LS GM (95% CI) for the Post/Pre Ratio<sup>2</sup> | 0.99 (0.79, 1.24) | 0.93 (0.73, 1.19) | 0.97 (0.76, 1.23) | 0.720 | 0.902 | 0.817 |

<sup>1</sup>Sample sizes were lower for Day 28 (n = 21 bLF; n = 18 low-dose rhLF; n = 20 high-dose rhLF); Day 56 (n = 20 bLF; n = 18 low-dose rhLF; n = 17 high-dose rhLF); and Day 84 (n = 20 bLF; n = 18 low-dose rhLF; n = 17 high-dose rhLF) LS GM (95% CI) were obtained from analysis of covariance (ANCOVA) models that included the natural ln-transformed changes from baseline (expressed as the post/pre ratio) as the dependent variable, product and sex as fixed effects, and the untransformed baseline as a
covariate. If non-normality of model residuals persisted with the ln transformations, a ranked model was also run. There were no material differences between the two models, so nominal p-values were reported from the ranked model, which were not adjusted for multiple comparisons or the testing of multiple study endpoints. All safety outcomes were tested at $\alpha = 0.05$ , 2-sided, significance level to minimize the risk of a type II error.
<sup>2</sup>Indicates that the ln-transformation resolved the non-normality, so the p-value was generated from the ln-transformed model.
<sup>3</sup>Indicates that the ANCOVA model included a significant ( $P < 0.05$ ) sex-by-product interaction term.
Abbreviations: bLF, bovine lactoferrin; hs-CRP, high-sensitivity C-reactive protein; IQRL, interquartile range limits; ln, natural log; LS GM, least squares geometric mean; rhLF, recombinant human lactoferrin.

Overall, bLF and rhLF were well tolerated at all concentrations tested in Study 1 and there were no product-related dropouts. Study product-emergent AEs are described in **Table 6**. All AEs were unrelated to the study product except low iron saturation in 3 participants (n =1 bLF; n = 2 low-dose rhLF) that was judged to be possibly or probably related to the study product. All iron-related laboratory values for these three participants with AEs possibly or probably related to the study products are provided in **Supplemental Table 7**.

**Table 6.** Treatment-emergent adverse events in Study 1.

| **Treatment Group** | **Description** | **Severity** | **Relationship** | **Action** | **Outcome<sup>1</sup>** |
| --- | --- | --- | --- | --- | --- |
| **Bovine Lactoferrin** |  |  |  |  |  |
|  | Iron Deficiency Anemia | Mild | Not Related | No Change | Not Yet Recovered |
|  | Low Iron Saturation | Severe | Probably | No Change | Not Yet Recovered |
|  | Elevated AST/ALT | Mild | Not Related | No Change | Unknown |
|  | Cough | Moderate | Not Related | No Change | Recovered Completely |
|  | Cough | Moderate | Not Related | No Change | Recovered Completely |
| **Low-Dose Recombinant Human Lactoferrin** |  |  |  |  |  |
|  | Low Iron Saturation | Mild | Possibly | No Change | Recovered Completely |
|  | Low Iron Saturation | Severe | Probably | No Change | Not Yet Recovered |
| **High-Dose Recombinant Human Lactoferrin** |  |  |  |  |  |
|  | Premenstrual Syndrome | Mild | Not Related | Stopped | Recovered Completely |
|  | Anemia | Severe | Not Related | No Change | Recovered Completely |
|  | Iron Deficiency | Mild | Not Related | No Change | Recovered Completely |
|  | Abdominal Surgery<sup>2</sup> | Severe | Not Related | No Change | Not Yet Recovered |
|  | Bladder Rupture | Severe | Not Related | No Change | Recovered Completely |
|  | URI | Moderate | Not Related | No Change | Recovered Completely |
|  | Flu | Moderate | Not Related | No Change | Recovered Completely |
|  | URI | Mild | Not Related | No Change | Recovered Completely |

Abbreviations: ALT, Alanine Aminotransferase; AST, Aspartate Aminotransferase; URI, upper respiratory infection. <sup>1</sup>Not yet recovered indicates that the adverse event was not resolved when the participant was last contacted. <sup>2</sup>Abdominal surgery was a serious adverse event that was related to a motor vehicle accident.

### 3.3 Dietary Intake

Select dietary intake data are presented in **Supplemental Tables 8-9** for the ITT population in Study 1 and the evaluable population in Study 2. In Study 1, reported total median energy intake was less than 2000 kcals/day for all treatment groups. Reported median iron intakes were 8.9 mg/d, 9.5 mg/d and 10.1 mg/d for bLF, low-dose rhLF, and high-dose rhLF, respectively, at baseline (Day 0); reported median iron intakes were 10.1 mg/d, 12.1 mg/d and 10.0 mg/d for bLF, low-dose rhLF, and high-dose rhLF, respectively, at Day 28. The nutrient analysis of the diet records did not include the study products. For Study 2, the dietary intake data for the evaluable population was similar to Study 1. The reported total median energy intake was 1467 kcals/d on Day 0 and 1637 kcals/d on Day 28. The reported median iron intakes were 8.3 mg/d and 7.9 mg/d on Day 0 and Day 28, respectively.

## 3. DISCUSSION

In this study, the alloimmunization potential of rhLF was assessed to address this key unanswered question on the safety of rhLF as a food ingredient. Dozens of previous studies on rhLF and bLF have consistently shown that LF is well tolerated and safe but has not shown to have pharmacologic activity when evaluated in large, well-powered, clinical trials **(Supplemental Table 10)**. Until now, immunogenicity testing has been primarily used for biologics and drug safety testing. However, many food ingredients are known to have functions that interact with the immune system and immunogenicity testing is a way to ensure the protein/food ingredient does not lead to a potentially adverse immune response. Specifically, for an exogenous human protein, such as rhLF, the potential for alloimmunization and the development of antibodies against the endogenous protein is important to evaluate.

To adequately assess the immunogenicity/alloimmunization potential of a recombinant human protein for use as a food ingredient, two expert panels recommended clinical evaluation to demonstrate reasonable certainty of no harm (21). In Study 1, anti-bLF antibodies increased with consumption of bLF, but there were no material changes in anti-hLF antibodies in the low- or high-dose rhLF groups. These data suggest low immunogenicity/alloimmunization potential for Helaina rhLF given no increase was measured in anti-hLF antibodies ∼28 days after the last exposure. The 28-day timepoint after the last exposure was recommended by the Expert Panel in 2023 (21), and align with FDA immunogenicity guidance of measuring antibodies approximately 30 days after the last oral exposure (27). The low-dose rhLF supplementation was selected near the 90^th^ percentile estimated daily intake (EDI) based on the intended use food categories of Helaina rhLF according to NHANES food consumption data that were provided by the National Center for Health Statistics 2017-2018 National Health and Nutrition Examination Surveys (NHANES) (28). The high-dose rhLF supplementation group was recommended by the 2023 expert panel as a 10-fold safety margin above the 90^th^ percentile EDI (21). The bLF dose level was matched to the high-dose rhLF. Importantly, the observational study that was run in parallel confirmed that the change and variability of anti-hLF antibodies were similar to what was observed in the low-dose and high-dose rhLF groups, supporting the conclusion that oral ingestion of rhLF did not lead to an immunogenic response to enhance anti-hLF antibody production. On the other hand, the anti-bLF antibodies in the observation study did not show the same increase from baseline as was measured at each timepoint in Study 1, suggesting that the anti-bLF antibodies were a response to bLF consumption in the affected participants.

Bovine LF is GRAS in the US and approved as a food ingredient in the EU for various intended uses and it is readily available in the food supply (29, 30). The clinical significance of the heightened serum anti-bLF antibody response measured in Study 1 is uncertain; however, no serious AEs were noted throughout the study, including the 8-week follow-up period, suggesting no clinically relevant outcome. This was the first study in humans assessing anti-bLF antibodies after oral exposure for 28 days. It is notable that 3.4 g/d that was supplemented in this study is substantially higher than the mean (39 mg/d) and 90^th^ percentile (80 mg/d) EDI of bLF based on 2007-2008 NHANES for the US population due to ingesting cow’s milk and/or its byproducts (13). As was observed in Study 2, anti-bLF antibodies did not show a significant change from baseline at 28, 56, or 84 days, indicating the anti-bLF antibodies in Study 1 were study-product emergent. Furthermore, the anti-bLF responses demonstrate that the duration of the study was sufficient to evaluate immunogenicity from oral ingestion through the detection of antibody response. Regarding novel rhLF as a food ingredient, it was critical from a safety perspective that there was no induction of serum anti-hLF antibody at both dose levels. Importantly, as neither dose (0.34 g/d and 3.4 g/d) of rhLF led to study product-emergent anti-hLF antibodies, there is reasonable certainty that Helaina rhLF would not pose an immunogenic risk over longer exposure periods at the intended use level.

All AEs in Study 1 were unrelated to the study product except for low iron saturation in 3 participants (n =1 bLF; n = 2 low-dose rhLF). Despite three of the AEs being classified as either “possibly” or “probably” related to the study product, it is worth noting that these participants had abnormal iron saturation levels at baseline. Furthermore, it is important to interpret the results for iron saturation in the context of all other iron-related outcomes. Hemoglobin and hematocrit, which are part of the CBC that is routinely measured, are most often used to diagnose iron deficiency anemia in a clinical setting (31). For two of the three participants with AEs of low iron saturation, results for hemoglobin, hematocrit, and iron were within the normal limits throughout the study. For the third participant, hemoglobin was within the normal range throughout the study, and iron and hematocrit were abnormal at baseline. All other AEs were categorized as unrelated to the study product.

In conclusion, the findings of this study answer a pivotal safety question raised by two expert panels (21) regarding immunogenicity of an orally ingested food ingredient and support the safety and tolerability of rhLF at an intake level of up to 3.4 g/d. These data suggest that there is low alloimmunization potential for rhLF at the intake level that was studied.

## Data sharing

Data described in the manuscript and analytical code will be made available upon reasonable request.

## Statement of author contributions

RDP: Conceptulization, resources, writing-review and editing, supervision; LLG: Conceptualization, writing-original draft, visualization, supervision, project administration; CGA: Conceptualization, supervision, project administration; MLW: Formal analysis; AJC: Conceptualization, resources, supervision; NPR: Methodology, formal analysis; KCM: Conceptualization, writing-review and editing; supervision, funding acquisition; CM: Conceptualization, resources methodology, writing-review and editing, supervision.

