## Supplementary Material for "A Randomized, Double-Blind, Controlled Trial to Assess the Effects of Lactoferrin at Two Doses vs. Active Control on Immunological and Safety Parameters in Healthy Adults"

| **Supplemental Table 1.** Nutrient content of one serving (6 g) of bovine lactoferrin and recombinant human lactoferrin products | | | |
| --- | --- | --- | --- |
|  | **bLF** | **Low-dose rhLF** | **High-dose rhLF** |
| Energy (kcals) | 21 | 21 | 17 |
| Carbohydrate (g) | 3.5 | 5.0 | 2.3 |
| Fat (g) | 0 | 0 | 0 |
| Protein (g) | 1.8 | 0.2 | 2.1 |
| Abbreviations: bLF, bovine lactoferrin; rhLF, recombinant human lactoferrin. | | | |

| **Supplemental Table 2.** Serum anti-lactoferrin antibodies by sex at baseline (Day 0), Day 28, Day 56, and Day 84 in the intention-to-treat population in study 1^1^ | | | | | | |
| --- | --- | --- | --- | --- | --- | --- |
|  | | | | **P-Values** | | |
|  | **bLF** | **Low-Dose rhLF** | **High-Dose rhLF** | **bLF vs.**  **Low-Dose rhLF** | **bLF vs.**  **High-Dose rhLF** | **Low-Dose**  **vs. High-Dose rhLF** |
| **Females: Anti-LF Antibodies in Serum^2^** | **n = 11** | **n = 10** | **n =11** |  |  |  |
| Day 0 Median (IQRL) | 747 (545, 917) | 641 (151, 1030) | 311 (144, 705) |  |  |  |
| Day 28 LS GM (95% CI) for the Post/Pre Ratio | 2.95 (1.87, 4.65) | 1.10 (0.69, 1.74) | 0.89 (0.56, 1.42) | **0.001** | **<0.001** | 0.377 |
| Day 56 LS GM (95% CI) for the Post/Pre Ratio | 3.50 (2.23, 5.49) | 1.07 (0.70, 1.65) | 1.04 (0.51, 2.14) | **0.001** | **0.001** | 0.499 |
| Day 84 LS GM (95% CI) for the Post/Pre Ratio | 3.18 (2.17, 4.68) | 1.01 (0.67, 1.52) | 0.97 (0.63, 1.49) | **0.006** | **0.002** | 0.639 |
| **Males: Anti-LF Antibodies in Serum^2^** | **n = 12** | **n = 11** | **n = 11** |  |  |  |
| Day 0 Median (IQRL) | 544 (346, 693) | 320 (118, 973) | 609 (225, 1525) |  |  |  |
| Day 28 LS GM (95% CI) for the Post/Pre Ratio | 2.05 (1.32, 3.19) | 1.01 (0.66, 1.56) | 0.97 (0.64, 1.47) | **0.013** | **0.001** | 0.738 |
| Day 56 LS GM (95% CI) for the Post/Pre Ratio | 2.63 (1.48, 4.67) | 1.08 (0.64, 1.80) | 0.98 (0.58, 1.66) | **0.013** | **<0.001** | 0.192 |
| Day 84 LS GM (95% CI) for the Post/Pre Ratio | 2.63 (1.60, 4.32) | 1.07 (0.64, 1.80) | 0.97 (0.58, 1.62) | **0.006** | **<0.001** | 0.215 |
| ^1^Missing data at Days 28 and 56 were estimated using multiple imputation. Therefore, sample sizes were lower for Day 84 (bLF: n = 10 females, n = 10 males; low-dose rhLF: n = 9 females, n = 9 males; high-dose rhLF: n = 8 females, n = 9 males). LS GM (95% CI) were obtained from analysis of covariance models that included the natural ln-transformed changes from baseline (expressed as the post/pre ratio) as the dependent variable, product as the fixed effect, and the untransformed baseline as a covariate. Non-normality of model residuals persisted with the ln transformations, so a ranked model was also run. There were no material differences between the two models, so nominal p-values are reported from the ranked model, which were not adjusted for multiple comparisons or the testing of multiple study endpoints. For the primary outcome (serum anti-LF antibodies at Day 56), differences between the product groups with each sex were tested using a hierarchical approach. Comparison of the high-dose rhLF group to the active control was tested at the α = 0.05, 2-sided, significance level. Comparison of the high-dose to the low-dose rhLF was tested at α = 0.0253, 2-sided, significance level. Comparison of the low-dose recombinant group to the active control was tested at α = 0.0167, 2-sided, significance level. Exploratory outcomes (serum anti-LF antibodies at Days 28 and 84) were tested at α = 0.05, 2-sided, significance level to minimize the risk of a type II error.  ^2^Serum anti-bLF antibodies were measured in the bLF group, and serum anti-human LF antibodies were measured in the low- and high-dose rhLF. Units for antibodies in serum are electrochemiluminescent (ECL) units.  Abbreviations: bLF, bovine lactoferrin; IQRL, interquartile range limits; ln, natural log; LS GM, least squares geometric mean; rhLF, recombinant human lactoferrin. | | | | | | |

| **Supplemental Table 3.** Serum anti-lactoferrin antibodies at baseline (Day 0), Day 28, Day 56, and Day 84 in the per protocol population in study 1^1^ | | | | | | |
| --- | --- | --- | --- | --- | --- | --- |
|  | | | | **P-Values** | | |
|  | **bLF** | **Low-Dose rhLF** | **High-Dose rhLF** | **bLF vs.**  **Low-Dose rhLF** | **bLF vs.**  **High-Dose rhLF** | **Low-Dose**  **vs. High-Dose rhLF** |
| **All Participants: Anti-LF Antibodies in Serum^3^** | **n = 19** | **n = 17** | **n = 15** |  |  |  |
| Day 0 Median (IQRL) | 585 (413, 787) | 320 (88.5, 959) | 417 (225, 1047) |  |  |  |
| Day 28 LS GM (95% CI) for the Post/Pre Ratio | 2.11 (1.59, 2.80) | 1.06 (0.79, 1.42) | 0.95 (0.69, 1.31) | **<0.001** | **<0.001** | 0.468 |
| Day 56 LS GM (95% CI) for the Post/Pre Ratio | 2.97 (2.14, 4.10) | 1.07 (0.76, 1.50) | 0.97 (0.67, 1.39) | **<0.001** | **<0.001** | 0.041 |
| Day 84 LS GM (95% CI) for the Post/Pre Ratio | 2.75 (2.02, 3.75) | 1.05 (0.76, 1.45) | 0.97 (0.69, 1.38) | **<0.001** | **<0.001** | 0.209 |
| **Females: Anti-LF Antibodies in Serum^3^** | **n = 9** | **n = 8** | **n = 6** |  |  |  |
| Day 0 Median (IQRL) | 747 (497, 816) | 438 (85.3, 983) | 313 (136, 391) |  |  |  |
| Day 28 LS GM (95% CI) for the Post/Pre Ratio^2^ | 2.36 (1.57, 3.53) | 1.13 (0.74, 1.73) | 0.92 (0.56, 1.53) | **0.017** | **0.007** | 0.523 |
| Day 56 LS GM (95% CI) for the Post/Pre Ratio | 3.14 (2.03, 4.88) | 1.08 (0.68, 1.71) | 0.97 (0.56, 1.68) | **0.008** | **0.002** | 0.321 |
| Day 84 LS GM (95% CI) for the Post/Pre Ratio^2^ | 2.86 (1.88, 4.34) | 1.03 (0.66, 1.61) | 0.97 (0.58, 1.62) | **0.003** | **0.003** | 0.852 |
| **Males: Anti-LF Antibodies in Serum^3^** | **n = 10** | **n = 9** | **n = 9** |  |  |  |
| Day 0 Median (IQRL) | 544 (378, 618) | 170 (110, 344) | 609 (257, 1434) |  |  |  |
| Day 28 LS GM (95% CI) for the Post/Pre Ratio | 1.93 (1.24, 2.99) | 0.99 (0.63, 1.57) | 0.93 (0.59, 1.46) | **0.011** | **0.001** | 0.431 |
| Day 56 LS GM (95% CI) for the Post/Pre Ratio | 2.82 (1.67, 4.76) | 1.07 (0.62, 1.83) | 0.95 (0.55, 1.62) | **0.007** | **<0.001** | 0.052 |
| Day 84 LS GM (95% CI) for the Post/Pre Ratio | 2.63 (1.60, 4.32) | 1.07 (0.64, 1.80) | 0.97 (0.58, 1.62) | **0.006** | **<0.001** | 0.215 |
| ^1^ LS GM (95% CI) were obtained from analysis of covariance models that included the natural ln-transformed changes from baseline (expressed as the post/pre ratio) as the dependent variable, product as the fixed effect, and the untransformed baseline as a covariate. Sex was also included as a fixed effect when the model was run in all participants. If non-normality of model residuals persisted with the ln-transformation, a ranked model was also run. There were no material differences between the two models, so nominal p-values are reported from the ranked model, which were not adjusted for multiple comparisons or the testing of multiple study endpoints. For the primary outcome (serum anti-LF antibodies at Day 56), differences between the product groups were tested within each sex and the overall sample using a hierarchical approach. Comparison of the high-dose rhLF group to the active control was tested at the α = 0.05, 2-sided, significance level. Comparison of the high-dose to the low-dose rhLF was tested at α = 0.0253, 2-sided, significance level. Comparison of the low-dose recombinant group to the active control was tested at α = 0.0167, 2-sided, significance level. Exploratory outcomes (serum anti-LF antibodies at Days 28 and 84) were tested at α = 0.05, 2-sided, significance level to minimize the risk of a type II error.  ^2^Indicates that the ln-transformation resolved the non-normality, so the p-values were generated from the ln-transformed model. ^3^Serum anti-bLF antibodies were measured in the bLF group, and serum anti-human LF antibodies were measured in the low- and high-dose rhLF. Units for antibodies in serum are electrochemiluminescent (ECL) units.  Abbreviations: bLF, bovine lactoferrin; IQRL, interquartile range limits; ln, natural log; LS GM, least squares geometric mean; rhLF, recombinant human lactoferrin. | | | | | | |

| **Supplemental Table 4.** Comprehensive metabolic panel at baseline (Day -7), Day 28, Day 56, and Day 84 in the safety population in Study 1^1^ | | | |
| --- | --- | --- | --- |
|  | **bLF**  **(n = 23)** | **Low-Dose rhLF**  **(n = 21)** | **High-Dose rhLF**  **(n = 22)** |
| **ALT (IU/L)** |  |  |  |
| Day -7 Median (IQRL) | 18.0 (12.0, 31.0) | 16.0 (14.0, 24.0) | 18.0 (14.0, 27.3) |
| Day 28 LS GM (95% CI) for the Post/Pre Ratio | 0.87 (0.76, 1.01) | 0.97 (0.83, 1.13) | 0.96 (0.83, 1.11) |
| Day 56 LS GM (95% CI) for the Post/Pre Ratio | 1.00 (0.86, 1.17) | 1.07 (0.91, 1.25) | 1.05 (0.89, 1.24) |
| Day 84 LS GM (95% CI) for the Post/Pre Ratio | 1.10 (0.93, 1.30) | 1.04 (0.87, 1.23) | 0.94 (0.79, 1.12) |
| **AST (IU/L)** |  |  |  |
| Day -7 Median (IQRL) | 20.0 (15.5, 23.0) | 18.0 (15.0, 22.0) | 18.0 (16.3, 23.0) |
| Day 28 LS GM (95% CI) for the Post/Pre Ratio | 0.95 (0.86, 1.05) | 0.91 (0.82, 1.01) | 1.00 (0.91, 1.11) |
| Day 56 LS GM (95% CI) for the Post/Pre Ratio | 1.05 (0.94, 1.18) | 1.01 (0.89, 1.14) | 1.10 (0.97, 1.25) |
| Day 84 LS GM (95% CI) for the Post/Pre Ratio | 1.15 (0.97, 1.37) | 1.02 (0.85, 1.23) | 0.99 (0.82, 1.20) |
| **Albumin (g/dL)** |  |  |  |
| Day -7 Median (IQRL) | 4.60 (4.40, 4.90) | 4.70 (4.40, 5.00) | 4.70 (4.50, 4.80) |
| Day 28 LS GM (95% CI) for the Post/Pre Ratio | 0.96 (0.93, 0.98) | 0.98 (0.96, 1.01) | 0.97 (0.95, 0.99) |
| Day 56 LS GM (95% CI) for the Post/Pre Ratio | 0.98 (0.95, 1.00) | 1.01 (0.98, 1.03) | 1.00 (0.97, 1.02) |
| Day 84 LS GM (95% CI) for the Post/Pre Ratio | 0.97 (0.94, 0.99) | 0.98 (0.95, 1.01) | 0.96 (0.93, 0.99) |
| **Alkaline Phosphatase (IU/L)** |  |  |  |
| Day -7 Median (IQRL) | 70.0 (62.0, 93.5) | 69.0 (57.0, 76.0) | 65.0 (52.5, 76.3) |
| Day 28 LS GM (95% CI) for the Post/Pre Ratio | 0.97 (0.92, 1.02) | 1.00 (0.95, 1.05) | 0.98 (0.94, 1.03) |
| Day 56 LS GM (95% CI) for the Post/Pre Ratio | 0.96 (0.92, 1.01) | 1.04 (0.98, 1.09) | 0.99 (0.94, 1.04) |
| Day 84 LS GM (95% CI) for the Post/Pre Ratio | 0.96 (0.92, 1.01) | 1.05 (1.00, 1.10) | 0.99 (0.94, 1.04) |
| **BUN (mg/dL)** |  |  |  |
| Day -7 Median (IQRL) | 11.0 (10.0, 13.0) | 12.0 (8.00, 13.0) | 12.5 (10.3, 16.0) |
| Day 28 LS GM (95% CI) for the Post/Pre Ratio | 1.04 (0.95, 1.14) | 1.12 (1.01, 1.24) | 1.04 (0.95, 1.14) |
| Day 56 LS GM (95% CI) for the Post/Pre Ratio | 0.99 (0.91, 1.08) | 1.00 (0.92, 1.10) | 1.06 (0.97, 1.17) |
| Day 84 LS GM (95% CI) for the Post/Pre Ratio | 1.01 (0.93, 1.10) | 1.01 (0.92, 1.11) | 0.94 (0.86, 1.04) |
| **Total Bilirubin (mg/dL)** |  |  |  |
| Day -7 Median (IQRL) | 0.40 (0.30, 0.55) | 0.40 (0.30, 0.60) | 0.40 (0.30, 0.58) |
| Day 28 LS GM (95% CI) for the Post/Pre Ratio | 0.78 (0.65, 0.94) | 0.82 (0.67, 0.99) | 0.79 (0.65, 0.94) |
| Day 56 LS GM (95% CI) for the Post/Pre Ratio | 0.89 (0.74, 1.06) | 0.85 (0.71, 1.03) | 0.92 (0.76, 1.11) |
| Day 84 LS GM (95% CI) for the Post/Pre Ratio | 1.04 (0.86, 1.26) | 0.93 (0.76, 1.14) | 0.90 (0.73, 1.11) |
| **Creatinine (mg/dL)** |  |  |  |
| Day -7 Median (IQRL) | 0.82 (0.65, 1.01) | 0.74 (0.70, 0.90) | 0.83 (0.67, 0.98) |
| Day 28 LS GM (95% CI) for the Post/Pre Ratio | 0.99 (0.96, 1.03) | 1.08 (1.03, 1.13) | 1.01 (0.97, 1.05) |
| Day 56 LS GM (95% CI) for the Post/Pre Ratio | 1.02 (0.97, 1.07) | 1.02 (0.97, 1.07) | 1.03 (0.98, 1.09) |
| Day 84 LS GM (95% CI) for the Post/Pre Ratio | 1.03 (0.99, 1.06) | 1.06 (1.02, 1.10) | 1.04 (1.00, 1.08) |
| **Glucose (mg/dL)** |  |  |  |
| Day -7 Median (IQRL) | 90.0 (87.0, 94.5) | 95.0 (89.0, 97.0) | 93.0 (88.0, 98.8) |
| Day 28 LS GM (95% CI) for the Post/Pre Ratio | 0.95 (0.89, 1.01) | 1.01 (0.94, 1.07) | 0.98 (0.92, 1.04) |
| Day 56 LS GM (95% CI) for the Post/Pre Ratio | 0.98 (0.95, 1.02) | 1.01 (0.98, 1.05) | 1.00 (0.97, 1.04) |
| Day 84 LS GM (95% CI) for the Post/Pre Ratio | 0.97 (0.92, 1.02) | 1.01 (0.96, 1.06) | 1.03 (0.98, 1.09) |
| **Sodium (mmol/L)** |  |  |  |
| Day -7 Median (IQRL) | 139 (138, 141) | 140 (138, 140) | 139 (137, 140) |
| Day 28 LS GM (95% CI) for the Post/Pre Ratio | 1.00 (0.99, 1.00) | 1.00 (0.99, 1.00) | 1.00 (0.99, 1.01) |
| Day 56 LS GM (95% CI) for the Post/Pre Ratio | 1.00 (1.00, 1.01) | 1.00 (0.99, 1.00) | 1.00 (0.99, 1.00) |
| Day 84 LS GM (95% CI) for the Post/Pre Ratio | 1.00 (0.99, 1.01) | 1.00 (0.99, 1.01) | 0.99 (0.99, 1.00) |
| **eGFR (mL/min/1.73)** |  |  |  |
| Day -7 Median (IQRL) | 110 (101, 116) | 114 (106, 117) | 110 (91.8, 116) |
| Day 28 LS GM (95% CI) for the Post/Pre Ratio | 1.01 (0.98, 1.05) | 0.94 (0.91, 0.98) | 1.01 (0.97, 1.04) |
| Day 56 LS GM (95% CI) for the Post/Pre Ratio | 1.01 (0.97, 1.05) | 0.97 (0.93, 1.01) | 1.00 (0.95, 1.04) |
| Day 84 LS GM (95% CI) for the Post/Pre Ratio | 0.99 (0.96, 1.03) | 0.95 (0.91, 0.98) | 0.98 (0.94, 1.02) |
| ^1^Sample sizes were lower for Day 28 (n = 21 bLF; n = 18 low-dose rhLF; n = 20 high-dose rhLF); Day 56 (n = 20 bLF; n = 18 low-dose rhLF; n = 17 high-dose rhLF); and Day 84 (n = 20 bLF; n = 18 low-dose rhLF; n = 17 high-dose rhLF). LS GM (95% CI) were obtained from analysis of covariance models that included the natural ln-transformed changes from baseline (expressed as the post/pre ratio) as the dependent variable, product and sex as fixed effects, and the untransformed baseline as a covariate. A separate model was run for each lab test and each post-baseline visit. Participants with infinite ratio change values (resulting from a baseline value of 0) were excluded from the model that generated the LS GMs. Abbreviations: ALT, alanine aminotransferase; AST, aspartate aminotransferase; bLF, bovine lactoferrin; BUN, blood urea nitrogen; eGFR, estimated glomerular filtration rate; IQRL, interquartile range limits; IU, international units; ln, natural log; LS GM, least squares geometric mean; rhLF, recombinant human lactoferrin | | | |

| **Supplemental Table 5.** Complete blood count with differential/platelet outcome measures at baseline (Day -7), Day 28, Day 56, and Day 84 in the safety population in Study 1^1^ | | | |
| --- | --- | --- | --- |
|  | **bLF**  **(n = 23)** | **Low-Dose rhLF**  **(n = 21)** | **High-Dose rhLF**  **(n = 22)** |
| **Hematocrit (%)** |  |  |  |
| Day -7 Median (IQRL) | 41.3 (39.5, 46.0) | 42.2 (37.5, 46.4) | 42.3 (40.5, 46.4) |
| Day 28 LS GM (95% CI) for the Post/Pre Ratio | 0.98 (0.95, 1.01) | 0.96 (0.93, 0.99) | 0.97 (0.94, 1.00) |
| Day 56 LS GM (95% CI) for the Post/Pre Ratio | 0.98 (0.95, 1.00) | 0.99 (0.96, 1.02) | 0.98 (0.96, 1.01) |
| Day 84 LS GM (95% CI) for the Post/Pre Ratio | 1.00 (0.97, 1.02) | 1.00 (0.97, 1.03) | 0.98 (0.95, 1.01) |
| **Hemoglobin (g/dL)** |  |  |  |
| Day -7 Median (IQRL) | 13.7 (12.9, 14.4) | 14.4 (11.6, 15.5) | 13.8 (13.2, 15.2) |
| Day 28 LS GM (95% CI) for the Post/Pre Ratio | 0.98 (0.95, 1.01) | 0.95 (0.92, 0.98) | 0.97 (0.94, 1.00) |
| Day 56 LS GM (95% CI) for the Post/Pre Ratio | 0.98 (0.95, 1.01) | 0.98 (0.95, 1.01) | 0.98 (0.95, 1.01) |
| Day 84 LS GM (95% CI) for the Post/Pre Ratio | 0.98 (0.96, 1.01) | 0.99 (0.96, 1.02) | 0.97 (0.94, 1.00) |
| **Lymphocytes, Absolute (x10^3^/µL)** |  |  |  |
| Day -7 Median (IQRL) | 2.10 (1.70, 2.75) | 1.90 (1.70, 2.70) | 2.05 (1.63, 2.38) |
| Day 28 LS GM (95% CI) for the Post/Pre Ratio | 0.98 (0.91, 1.07) | 0.97 (0.89, 1.06) | 1.04 (0.96, 1.13) |
| Day 56 LS GM (95% CI) for the Post/Pre Ratio | 0.93 (0.84, 1.03) | 0.95 (0.86, 1.05) | 1.03 (0.93, 1.15) |
| Day 84 LS GM (95% CI) for the Post/Pre Ratio | 0.93 (0.85, 1.01) | 0.84 (0.76, 0.92) | 0.96 (0.87, 1.06) |
| **MCH (picograms)** |  |  |  |
| Day -7 Median (IQRL) | 29.1 (27.0, 30.4) | 29.3 (27.2, 30.6) | 29.2 (27.8, 30.3) |
| Day 28 LS GM (95% CI) for the Post/Pre Ratio | 1.00 (0.99, 1.00) | 0.99 (0.98, 0.99) | 1.00 (0.99, 1.01) |
| Day 56 LS GM (95% CI) for the Post/Pre Ratio | 0.99 (0.98, 1.01) | 0.99 (0.98, 1.00) | 0.99 (0.98, 1.00) |
| Day 84 LS GM (95% CI) for the Post/Pre Ratio | 0.98 (0.96, 0.99) | 0.98 (0.97, 1.00) | 0.99 (0.97, 1.00) |
| **MCHC (g/dL)** |  |  |  |
| Day -7 Median (IQRL) | 33.3 (31.9, 34.0) | 33.4 (32.4, 34.0) | 33.1 (32.3, 33.7) |
| Day 28 LS GM (95% CI) for the Post/Pre Ratio | 1.00 (0.99, 1.01) | 0.99 (0.98, 1.00) | 1.00 (0.99, 1.01) |
| Day 56 LS GM (95% CI) for the Post/Pre Ratio | 1.00 (0.99, 1.01) | 0.99 (0.98, 1.00) | 1.00 (0.99, 1.01) |
| Day 84 LS GM (95% CI) for the Post/Pre Ratio | 0.99 (0.98, 1.00) | 0.99 (0.98, 1.00) | 0.99 (0.98, 1.00) |
| **MCV (fL)** |  |  |  |
| Day -7 Median (IQRL) | 87.0 (83.5, 90.0) | 88.0 (84.0, 91.0) | 88.5 (83.0, 91.0) |
| Day 28 LS GM (95% CI) for the Post/Pre Ratio | 1.00 (0.99, 1.01) | 1.00 (0.99, 1.01) | 1.00 (0.99, 1.01) |
| Day 56 LS GM (95% CI) for the Post/Pre Ratio | 0.99 (0.98, 1.00) | 1.00 (0.99, 1.01) | 1.00 (0.98, 1.01) |
| Day 84 LS GM (95% CI) for the Post/Pre Ratio | 0.99 (0.98, 1.00) | 1.00 (0.98, 1.01) | 1.00 (0.99, 1.01) |
| **Monocytes, Absolute (x10^3^/µL)** |  |  |  |
| Day -7 Median (IQRL) | 0.50 (0.40, 0.55) | 0.50 (0.40, 0.60) | 0.50 (0.40, 0.60) |
| Day 28 LS GM (95% CI) for the Post/Pre Ratio | 1.03 (0.94, 1.14) | 0.99 (0.89, 1.10) | 1.07 (0.97, 1.18) |
| Day 56 LS GM (95% CI) for the Post/Pre Ratio | 0.94 (0.84, 1.05) | 0.91 (0.80, 1.02) | 0.92 (0.81, 1.04) |
| Day 84 LS GM (95% CI) for the Post/Pre Ratio | 0.95 (0.85, 1.05) | 1.02 (0.91, 1.14) | 1.00 (0.89, 1.13) |
| **Neutrophils, Absolute (x10^3^/µL)** |  |  |  |
| Day -7 Median (IQRL) | 3.90 (2.85, 4.60) | 3.00 (2.60, 3.50) | 3.35 (2.63, 4.03) |
| Day 28 LS GM (95% CI) for the Post/Pre Ratio | 1.01 (0.92, 1.11) | 1.00 (0.90, 1.11) | 1.01 (0.92, 1.11) |
| Day 56 LS GM (95% CI) for the Post/Pre Ratio | 0.92 (0.81, 1.05) | 1.02 (0.88, 1.17) | 0.99 (0.86, 1.14) |
| Day 84 LS GM (95% CI) for the Post/Pre Ratio | 0.88 (0.78, 1.00) | 1.02 (0.89, 1.16) | 0.98 (0.86, 1.12) |
| **Platelets (x10^3^/µL)** |  |  |  |
| Day -7 Median (IQRL) | 260 (236, 304) | 283 (229, 336) | 261 (226, 297) |
| Day 28 LS GM (95% CI) for the Post/Pre Ratio | 0.99 (0.94, 1.05) | 1.03 (0.97, 1.09) | 0.97 (0.92, 1.03) |
| Day 56 LS GM (95% CI) for the Post/Pre Ratio | 0.98 (0.93, 1.03) | 1.07 (1.02, 1.13) | 1.04 (0.98, 1.10) |
| Day 84 LS GM (95% CI) for the Post/Pre Ratio | 1.00 (0.95, 1.05) | 1.02 (0.97, 1.08) | 1.02 (0.96, 1.08) |
| **RBC (x10^6^/µL)** |  |  |  |
| Day -7 Median (IQRL) | 4.75 (4.40, 5.40) | 4.75 (4.45, 5.21) | 4.85 (4.51, 5.44) |
| Day 28 LS GM (95% CI) for the Post/Pre Ratio | 0.98 (0.96, 1.01) | 0.96 (0.94, 0.99) | 0.97 (0.95, 1.00) |
| Day 56 LS GM (95% CI) for the Post/Pre Ratio | 0.99 (0.96, 1.01) | 0.99 (0.97, 1.02) | 0.99 (0.96, 1.02) |
| Day 84 LS GM (95% CI) for the Post/Pre Ratio | 1.01 (0.98, 1.03) | 1.01 (0.98, 1.03) | 0.98 (0.96, 1.01) |
| **RDW (%)** |  |  |  |
| Day -7 Median (IQRL) | 12.9 (12.7, 13.7) | 13.2 (12.5, 13.6) | 13.0 (12.6, 13.4) |
| Day 28 LS GM (95% CI) for the Post/Pre Ratio | 0.99 (0.97, 1.01) | 1.01 (0.99, 1.04) | 1.00 (0.98, 1.02) |
| Day 56 LS GM (95% CI) for the Post/Pre Ratio | 0.99 (0.96, 1.02) | 1.01 (0.98, 1.04) | 0.99 (0.96, 1.03) |
| Day 84 LS GM (95% CI) for the Post/Pre Ratio | 0.99 (0.97, 1.01) | 1.01 (0.98, 1.04) | 0.98 (0.96, 1.01) |
| **WBC (x10^3^/µL)** |  |  |  |
| Day -7 Median (IQRL) | 6.80 (5.25, 7.75) | 5.70 (5.00, 6.70) | 6.15 (5.03, 7.05) |
| Day 28 LS GM (95% CI) for the Post/Pre Ratio | 1.00 (0.93, 1.08) | 0.99 (0.92, 1.08) | 1.03 (0.96, 1.11) |
| Day 56 LS GM (95% CI) for the Post/Pre Ratio | 0.94 (0.86, 1.03) | 0.97 (0.89, 1.07) | 1.02 (0.93, 1.12) |
| Day 84 LS GM (95% CI) for the Post/Pre Ratio | 0.92 (0.84, 1.00) | 0.94 (0.86, 1.03) | 1.00 (0.91, 1.10) |
| ^1^Sample sizes were lower for Day 28 (n = 21 bLF; n = 18 low-dose rhLF; n = 20 high-dose rhLF); Day 56 (n = 20 bLF; n = 18 low-dose rhLF; n = 17 high-dose rhLF); and Day 84 (n = 20 bLF; n = 18 low-dose rhLF; n = 17 high-dose rhLF). LS GM (95% CI) were obtained from analysis of covariance models that included the natural ln-transformed changes from baseline (expressed as the post/pre ratio) as the dependent variable, product and sex as fixed effects, and the untransformed baseline as a covariate. A separate model was run for each lab test and each post-baseline visit. Participants with infinite ratio change values (resulting from a baseline value of 0) were excluded from the model that generated the LS GMs. Abbreviations: bLF, bovine lactoferrin; fL, femtoliters; IQRL, interquartile range limits; ln, natural log; LS GM, least squares geometric mean; MCH, mean corpuscular hemoglobin; MCHC, mean corpuscular hemoglobin concentration; MCV, mean corpuscular volume; RBC, red blood cell; RDW, red cell distribution width; WBC, white blood cell; rhLF, recombinant human lactoferrin. | | | |

| **Supplemental Table 6.** Urinalysis continuous outcome measures at baseline (Day -7), Day 28, Day 56, and Day 84 in the safety population in study 1^1^ | | | |
| --- | --- | --- | --- |
|  | **bLF**  **(n = 23)** | **Low-Dose rhLF**  **(n = 21)** | **High-Dose rhLF**  **(n = 22)** |
| **Specific Gravity** |  |  |  |
| Day -7 Median (IQRL) | 1.02 (1.02, 1.03) | 1.02 (1.02, 1.03) | 1.02 (1.01, 1.02) |
| Day 28 LS GM (95% CI) for the Post/Pre Ratio | 1.02 (1.01, 1.02) | 1.01 (1.01, 1.02) | 1.01 (1.01, 1.02) |
| Day 56 LS GM (95% CI) for the Post/Pre Ratio | 1.02 (0.96, 1.08) | 0.97 (0.91, 1.04) | 0.97 (0.91, 1.04) |
| Day 84 LS GM (95% CI) for the Post/Pre Ratio | 1.02 (1.01, 1.02) | 1.01 (1.01, 1.02) | 1.01 (1.01, 1.02) |
| **pH** |  |  |  |
| Day -7 Median (IQRL) | 6.00 (6.00, 7.00) | 6.00 (6.00, 6.50) | 6.75 (6.00, 7.00) |
| Day 28 LS GM (95% CI) for the Post/Pre Ratio | 0.96 (0.93, 0.99) | 0.97 (0.93, 1.00) | 0.96 (0.93, 1.00) |
| Day 56 LS GM (95% CI) for the Post/Pre Ratio | 0.99 (0.96, 1.03) | 1.02 (0.98, 1.05) | 0.98 (0.94, 1.01) |
| Day 84 LS GM (95% CI) for the Post/Pre Ratio | 0.98 (0.95, 1.02) | 0.97 (0.93, 1.01) | 0.95 (0.91, 0.99) |
| ^1^Sample sizes were lower for Day 28 (n = 21 bLF; n = 18 low-dose rhLF; n = 20 high-dose rhLF); Day 56 (n = 20 bLF; n = 18 low-dose rhLF; n = 17 high-dose rhLF); and Day 84 (n = 20 bLF; n = 18 low-dose rhLF; n = 17 high-dose rhLF). LS GM (95% CI) were obtained from analysis of covariance models that included the natural ln-transformed changes from baseline (expressed as the post/pre ratio) as the dependent variable, product and sex as fixed effects, and the untransformed baseline as a covariate. A separate model was run for each lab test and each post-baseline visit. Participants with infinite ratio change values (resulting from a baseline value of 0) were excluded from the model that generated the LS GMs. Abbreviations: bLF, bovine lactoferrin; IQRL, interquartile range limits; ln, natural log; LS GM, least squares geometric mean; rhLF, recombinant human lactoferrin. | | | |

| **Supplemental Table 7.** Iron-related laboratory values for participants with AEs of low iron saturation that were possibly or probably related to study products | | | | | |
| --- | --- | --- | --- | --- | --- |
|  | **Baseline (Day -7 or 0)** | **Visit 4 (Day 28)** | **Visit 6 (Day 56)** | **Visit 7 (Day 84)** | **Reference Range** |
| **Participant 107: AE of low iron saturation (mild severity; possibly related to low-dose rhLF)** | | | | | |
| Iron Saturation (%) | 13 | 10 | 7^1^ | 18^2^ | 15 - 55 |
| Iron (µg/dL) | 69 | 53 | 34 | 93 | 27 - 159 |
| Hemoglobin (g/dL) | 11.5 | 11.9 | 11.3 | 11.6 | 11.1 - 15.9 |
| Hematocrit (%) | 37.0 | 37.2 | 36.0 | 37.1 | 34.0 - 46.6 |
| Ferritin (ng/mL) | 19 | 33 | 20 | 36 | 30 – 400 |
| sTfR (nmol/L) | 27.4 | 23.5 | 22.0 | 22.8 | 12.2 – 27.3 |
| Total Iron Binding Capacity (µg/dL) | 533 | 547 | 481 | 511 | 250 - 450 |
| UIBC (µg/dL) | 464 | 494 | 447 | 418 | 131 - 425 |
| *Visit Dates* | *9/12/2023, 9/20/2023* | *10/17/2023* | *11/15/2023* | *12/13/2023* |  |
| *First/Last Day of Last Menstruation* | *8/15 – 8/30/2023* | *9/15 – 9/20/2023* | *10/9 – 10/17/2023* | *11/15 – 11/19/2023* |  |
| *Relevant Concomitant Medications: Ferrous sulfate (325 mg/d) was started 11/22/2023 and ongoing at the end of the study.* | | | | | |
| **Participant 112: AE of low iron saturation (severe severity; probably related to bLF)** | | | | | |
| Iron Saturation (%) | 12 | 8^1^ | 8 | 10^3^ | 15 - 55 |
| Iron (µg/dL) | 53 | 37 | 39 | 47 | 27 - 159 |
| Hemoglobin (g/dL) | 12.4 | 11.9 | 12.7 | 12.7 | 11.1 - 15.9 |
| Hematocrit (%) | 39.3 | 37.3 | 40.0 | 40.8 | 34.0 - 46.6 |
| Ferritin (ng/mL) | 41 | 17 | 23 | 39 | 30 – 400 |
| sTfR (nmol/L) | 25.4 | 23.2 | 19 | 17 | 12.2 – 27.3 |
| Total Iron Binding Capacity (µg/dL) | 460 | 445 | 469 | 460 | 250 - 450 |
| UIBC (µg/dL) | 407 | 408 | 430 | 413 | 131 - 425 |
| *Visit Date* | *9/12/2023, 9/20/2023* | *10/18/2023* | *11/15/2023* | *12/13/2023* |  |
| *First/Last Day of Last Menstruation* | *2/26 – 2/28/2023* | *2/26 – 2/28/2023* | *2/26 – 2/28/2023* | *2/28 – 3/2/2023* |  |
| *Relevant Concomitant Medications: None* | | | | | |
| **Participant 141: AE of low iron saturation (severe severity; probably related to low-dose rhLF)** | | | | | |
| Iron Saturation (%) | 77 | 11 | 9^1^ | 95^3^ | 15 - 55 |
| Iron (µg/dL) | 236 | 38 | 32 | 319 | 27 - 159 |
| Hemoglobin (g/dL) | 11.1 | 10.8 | 10.5 | 11.5 | 11.1 - 15.9 |
| Hematocrit (%) | 33.6 | 33.7 | 32.3 | 35.3 | 34.0 - 46.6 |
| Ferritin (ng/mL) | 37 | 13 | 12 | 50 | 30 – 400 |
| sTfR (nmol/L) | 23.3 | 19.8 | 26.5 | 25.1 | 12.2 – 27.3 |
| Total Iron Binding Capacity (µg/dL) | 306 | 333 | 352 | 337 | 250 - 450 |
| UIBC (µg/dL) | 70 | 295 | 320 | 18 | 131 - 425 |
| *Visit Date* | *9/15/2023, 9/21/2023* | *10/19/2023* | *11/16/2023* | *12/14/2023* |  |
| *First/Last Day of Last Menstruation* | *9/11 – 9/15/2023* | *10/10 -10/14/2023* | *11/10 – 11/14/2023* | *12/10 – 12/14/2023* |  |
| *Relevant Concomitant Medications: None* | | | | | |
| Values in red font indicate abnormal values. Baseline hemoglobin and hematocrit were measured on Day -7, and all other baseline iron-related labs were measured on Day 0. Abbreviations: AE, adverse event; bLF, bovine lactoferrin; rhLF, recombinant human lactoferrin; sTfR, soluble transferrin receptor; UIBC, unsaturated iron binding capacity. ^1^Indicates the laboratory value that corresponds to the start of the AE. ^2^Indicates the laboratory value that corresponds to the stop of the AE. ^3^Indicates that the AE was ongoing at the end of the study. | | | | | |

| **Supplemental Table 8.** Select nutrient measures at baseline (Day 0) and Day 28 in the intent-to-treat population in Study 1^1^ | | | |
| --- | --- | --- | --- |
|  | **bLF**  **(n = 22)** | **Low-Dose rhLF**  **(n = 20)** | **High-Dose rhLF**  **(n = 22)** |
| **Calories** |  |  |  |
| Day 0 Median (IQRL) | 1646 (968, 1935) | 1580 (1049, 2227) | 1702 (1469, 2032) |
| Day 28 Median (IQRL) | 1679 (1321, 2116) | 1908 (1558, 2619) | 1613 (1156, 1913) |
| Median Percent Change (IQRL) | 10.8 (-29.9, 64.8) | 14.4 (-10.3, 63.5) | 5.39 (-25.3, 16.2) |
| **Total Fat (g)** |  |  |  |
| Day 0 Median (IQRL) | 62.0 (38.2, 94.1) | 56.7 (37.6, 83.7) | 75.7 (56.8, 83.8) |
| Day 28 Median (IQRL) | 66.3 (53.8, 97.3) | 85.9 (57.4, 105) | 67.4 (54.8, 90.6) |
| Median Percent Change (IQRL) | 9.77 (-19.6, 87.9) | 35.3 (11.1, 118) | -1.15 (-16.8, 27.9) |
| **Saturated Fats (g)** |  |  |  |
| Day 0 Median (IQRL) | 17.4 (11.5, 30.1) | 17.5 (8.99, 25.8) | 22.9 (16.8, 27.5) |
| Day 28 Median (IQRL) | 20.1 (15.5, 29.5) | 26.4 (17.7, 34.9) | 22.1 (14.2, 30.8) |
| Median Percent Change (IQRL) | 9.53 (-14.1, 89.4) | 31.6 (-3.48, 100) | 4.48 (-11.0, 21.1) |
| **Polyunsaturated Fats (g)** |  |  |  |
| Day 0 Median (IQRL) | 9.26 (5.79, 13.4) | 8.87 (6.13, 13.0) | 11.1 (6.27, 15.2) |
| Day 28 Median (IQRL) | 11.1 (7.30, 14.3) | 13.5 (8.72, 17.1) | 8.44 (6.45, 11.4) |
| Median Percent Change (IQRL) | 0.42 (-37.4, 66.7) | 45.4 (11.2, 62.4) | -0.72 (-33.8, 91.5) |
| **Monounsaturated Fats (g)** |  |  |  |
| Day 0 Median (IQRL) | 16.4 (6.77, 22.0) | 13.6 (9.22, 24.1) | 18.1 (10.6, 23.4) |
| Day 28 Median (IQRL) | 19.0 (13.4, 28.8) | 20.8 (15.0, 33.1) | 19.8 (14.6, 28.7) |
| Median Percent Change (IQRL) | 7.89 (-39.0, 156) | 40.2 (-0.75, 110) | 9.25 (-24.3, 111) |
| **Protein (g)** |  |  |  |
| Day 0 Median (IQRL) | 66.0 (39.8, 92.9) | 69.3 (49.0, 86.3) | 80.0 (65.4, 161) |
| Day 28 Median (IQRL) | 70.8 (59.8, 103) | 90.8 (72.6, 107) | 79.8 (50.8, 97.0) |
| Median Percent Change (IQRL) | 28.9 (-15.7, 91.2) | 72.5 (-11.8, 93.8) | 4.81 (-29.9, 41.1) |
| **Total Carbohydrates (g)** |  |  |  |
| Day 0 Median (IQRL) | 181 (139, 235) | 184 (126, 236) | 151 (124, 207) |
| Day 28 Median (IQRL) | 181 (154, 225) | 196 (168, 266) | 168 (115, 202) |
| Median Percent Change (IQRL) | 12.4 (-26.9, 66.0) | 13.3 (-9.12, 78.1) | 10.7 (-30.0, 29.7) |
| **Sugars (g)** |  |  |  |
| Day 0 Median (IQRL) | 55.0 (26.6, 78.0) | 49.1 (18.9, 65.4) | 54.3 (27.8, 66.8) |
| Day 28 Median (IQRL) | 67.7 (53.7, 84.9) | 58.6 (37.8, 84.5) | 46.7 (22.1, 78.5) |
| Median Percent Change (IQRL) | 17.0 (2.94, 156) | 27.4 (-18.2, 158) | -14.2 (-31.5, 13.7) |
| **Fiber (g)** |  |  |  |
| Day 0 Median (IQRL) | 9.45 (6.85, 15.8) | 10.4 (7.96, 15.8) | 11.3 (7.63, 17.8) |
| Day 28 Median (IQRL) | 11.5 (7.31, 13.9) | 14.4 (10.3, 20.2) | 10.1 (8.09, 18.5) |
| Median Percent Change (IQRL) | 8.10 (-38.7, 53.3) | 23.3 (-11.5, 70.2) | 16.7 (-22.3, 55.0) |
| **Iron (mg)** |  |  |  |
| Day 0 Median (IQRL) | 8.90 (5.86, 11.7) | 9.53 (6.31, 12.3) | 10.1 (7.15, 13.4) |
| Day 28 Median (IQRL) | 10.1 (8.43, 11.8) | 12.1 (9.40, 14.3) | 9.97 (6.11, 12.7) |
| Median Percent Change (IQRL) | 26.0 (-11.9, 49.5) | 20.9 (11.6, 83.6) | 15.7 (-2.72, 48.1) |
| ^1^Sample sizes were lower for Day 28 (n = 22 bLF; n = 18 low-dose rhLF; n = 20 high-dose rhLF). Data are the averages from the 3-day diet records. If fewer than 3 days were completed, the average of the available days were used. Percent change computations that resulted in an infinite or undefined value were set to missing. Baseline diet records were missing for two participants in the intention-to-treat population. The study product was not included in the nutrient analysis of the diet records. | | | |

| **Supplemental Table 9.** Select nutrient measures at baseline (Day 0) and Day 28 in the evaluable population in Study 2^1^ | |
| --- | --- |
|  | **Evaluable Population**  **(N = 24)** |
| **Calories** |  |
| Day 0 Median (IQRL) | 1467 (1239, 1817) |
| Day 28 Median (IQRL) | 1637 (1342, 2081) |
| Median Percent Change (IQRL) | 8.47 (0.39, 22.9) |
| **Total Fat (g)** |  |
| Day 0 Median (IQRL) | 59.5 (51.5, 77.9) |
| Day 28 Median (IQRL) | 70.4 (53.8, 92.2) |
| Median Percent Change (IQRL) | 7.62 (-11.5, 48.2) |
| **Saturated Fats (g)** |  |
| Day 0 Median (IQRL) | 19.9 (15.4, 24.8) |
| Day 28 Median (IQRL) | 18.7 (15.5, 31.3) |
| Median Percent Change (IQRL) | 6.53 (-10.6, 40.2) |
| **Polyunsaturated Fats (g)** |  |
| Day 0 Median (IQRL) | 8.59 (5.82, 11.0) |
| Day 28 Median (IQRL) | 12.8 (9.76, 16.4) |
| Median Percent Change (IQRL) | 53.3 (-23.5, 152) |
| **Monounsaturated Fats (g)** |  |
| Day 0 Median (IQRL) | 14.5 (10.8, 23.6) |
| Day 28 Median (IQRL) | 17.9 (12.7, 27.2) |
| Median Percent Change (IQRL) | 24.6 (-17.7, 64.8) |
| **Protein (g)** |  |
| Day 0 Median (IQRL) | 60.9 (50.3, 99.6) |
| Day 28 Median (IQRL) | 67.4 (48.8, 105) |
| Median Percent Change (IQRL) | 2.65 (-17.5, 31.1) |
| **Total Carbohydrates (g)** |  |
| Day 0 Median (IQRL) | 156 (146, 184) |
| Day 28 Median (IQRL) | 174 (157, 215) |
| Median Percent Change (IQRL) | 10.5 (-6.73, 32.1) |
| **Sugars (g)** |  |
| Day 0 Median (IQRL) | 49.0 (29.3, 63.7) |
| Day 28 Median (IQRL) | 56.7 (44.1, 70.3) |
| Median Percent Change (IQRL) | 14.6 (-14.1, 55.6) |
| **Fiber (g)** |  |
| Day 0 Median (IQRL) | 10.2 (8.69, 11.7) |
| Day 28 Median (IQRL) | 10.3 (6.12, 14.1) |
| Median Percent Change (IQRL) | 7.50 (-38.7, 54.0) |
| **Iron (mg)** |  |
| Day 0 Median (IQRL) | 8.30 (5.80, 9.93) |
| Day 28 Median (IQRL) | 7.89 (6.34, 9.79) |
| Median Percent Change (IQRL) | -12.8 (-23.3, 50.5) |
| ^1^Data are the averages from the 3-day diet records. If fewer than 3 days were completed, the average of the available days were used. Percent change computations that resulted in an infinite or undefined value were set to missing. | |

| Supplemental Table 10. Clinical Trials of bLf and rhLf Exploring Safety and Pharmacologic Activities*  (*bold studies are with hLF from a bioengineered source and non-bold studies are bLF) | | | |
| --- | --- | --- | --- |
| **Reference** | **Study Design** | **Safety** | **Efficacy** |
| **Iron Status** | | | |
| (Abu Hashim et al., 2017) | Systematic Review and Meta Analysis: Iron deficiency anemia (IDA) during pregnancy. Primary outcome change in hemoglobin level at 4 weeks of treatment. 4 eligible trials (600 women) were analyzed. | Significantly less gastrointestinal side effects were reported with lactoferrin treatment than with oral ferrous iron preparations. | For pregnant women with IDA, daily oral treatment with bLf is just as good as ferrous sulfate in increasing Hb and other hematological parameters |
| (Chen et al., 2020) | Randomized, controlled, open and post-market intervention study of 108 anemic infants aged 6-9 months. bLf concentration was 0 mg/100 g, 38 mg/100 g, 76 mg/100 g formula; intervention was for 3 months. Daily intake was 0, 47.2 ± 8.8 mg and 91.5 ± 12.5 mg, respectively. | No adverse effects related to bLf administration were reported; no adverse effects on anthropometric indices. | No difference in Hb levels with bLf fortified infant formula at 1 month compared to control; after 3 months, the higher dose of bLf (76 mg/100 g formula compared to 38 mg/100 g formula) significantly increased Hb levels of anemic infants. |
| (El Amrousy et al., 2022) | Randomized clinical trial of 80 children with inflammatory bowel disease related iron-deficiency anemia given either ferrous sulfate at 6 mg/kg/day or lactoferrin at 100 mg/day for 3 months. | Lactoferrin was well tolerated with fewer gastrointestinal side effects than ferrous sulfate (reported in only one patient in the bLf group and 18 patients in the ferrous sulfate group). | Lactoferrin significantly increased Hb, serum iron, transferrin saturation, and serum ferritin compared to ferrous sulfate. |
| (Lepanto et al., 2018) | Interventional study was conducted by orally administering 100mg of 20–30% iron-saturated bLf (corresponding to 70–84 μg of elemental iron) twice a day for 30 days. This treatment was compared with the Italian standard therapy, consisting in the oral administration of 329.7mg of ferrous sulfate once a day (corresponding to 105mg of elemental iron). Treatments were carried out on 29 anemic women with minor b-thalassemia (20 pregnant and 9 non-pregnant), 149 women with hereditary thrombophilia (HT) (70 pregnant and 79 non-pregnant) affected by anemia of inflammation (AI) and 20 anemic pregnant women suffering from various pathologies. | The adverse effects of bLf and ferrous sulfate treatments on fetuses were monitored through ultrasonographic measurements of intrauterine growth and by the detection of amniotic fluid amount, expressed as AFI (39). No growth restriction was observed for any fetus. The adverse effects on new-born infants were checked by weight and APGAR score values which were not adversely affected. | In minor b-thalassemic pregnant women treated for 30 days with bLf a significant increase of Hb as well as a significant decrease of IL-6 was noted. bLf was more efficient than ferrous sulfate in AI treatment in HT pregnant and non-pregnant women by decreasing both serum IL-6 and hepcidin, thus increasing hematological parameters, such as the number of red blood cells (RBCs), the concentration of hemoglobin, total serum iron and serum ferritin. |
| (Nappi et al., 2009) | A prospective, randomized, controlled, double blind trial of 100 pregnant women treated either with one capsule of 100 mg bLf twice a day or 520 mg ferrous sulfate once a day for 30 days. | bLf was well tolerated in comparison to ferrous sulfate; median scores of abdominal pain and constipation were significantly higher in subject treated with ferrous sulfate in comparison with those treated with bLf. | In both groups, hemoglobin, serum ferritin and iron were significantly increased while total iron-binding capacity was significantly reduced in comparison with basal values; there were no significant differences between groups. |
| (Paesano et al., 2006) | bLf was given orally twice a day in a tablet containing 100 mg to pregnant women with iron deficiency or iron deficiency anemia for 30 days (n=107). Positive controls received 520 mg of ferrous sulfate administered orally once a day (n = 98). The dose of ferrous sulfate corresponded to 156 mg elemental iron and 200 mg bLf corresponded to 8.8 mg ferric ions. The control group did not receive any iron supplementation (n=54) | No adverse effects related to bLf administration were reported. | Total serum iron values were significantly higher in bLf treated women than in the ferrous sulfate treated group. Hemoglobin and total serum iron values were higher in both treated groups than the controls. Hemoglobin values were not significantly different in the bLf group compared to the ferrous sulfate treated group. |
| (Paesano et al., 2012) | Open-label cohort study in iron deficient and iron deficient anemic pregnant women given 100 mg bLf b.i.d. for at least 4 weeks until delivery. 163 women were enrolled. | No adverse side effects of bLf oral administration were noted as monitored through the following parameters: gastrointestinal discomfort, nausea, vomiting, diarrhea, and constipation. In addition, the following clinical laboratory parameters were evaluated every 30 days: hematocrit, glycemia, uricemia, bilirubin, glutamic oxaloacetic transaminase, glutamic pyruvic transaminase, cholesterol, triglyceride acid, and electrolytes. | A significant improvement of hematological parameters including red blood cell number, hemoglobin, total serum iron, and serum ferritin concentrations was observed in the women’s cohort orally receiving bLf, together with a consistent decrease in serum IL-6 levels. |
| (Paesano et al., 2014) | Interventional study evaluating the safety and efficacy of bLf, versus the ferrous sulphate standard intervention, in 295 pregnant women with iron deficiency and iron deficiency anemia affected by hereditary thrombophilia. 156 women received 100 mg bLf b.i.d. and 139 women received 520 mg ferrous sulfate once a day. Dosing lasted until delivery. | No adverse effects related to bLf administration were reported. | Red blood cells, hemoglobin, total serum iron, serum ferritin (hematological parameters) were assayed before and every 30 days during therapy until delivery and were significantly higher in women given bLf compared to ferrous sulfate. Serum IL-6 was measured at enrollment and after therapy at delivery and was significantly decreased in bLf treated women and increased in ferrous sulfate treated women. |
| (Rosa et al., 2020) | Clinical trial of 35 women where bLf (100 mg two times/day) was orally administered for 30 days before (Arm A n=17) or during meals (Arm B n=14) to pregnant women with hereditary thrombophilia and suffering from anemia of inflammation. | No adverse effects related to bLf administration were reported. | A significant increase of the number of red blood cells (RBCs), hemoglobin (Hb), total serum iron (TSI) and serum ferritin (sFtn) levels, along with a significant decrease of IL-6 were detected after 30 days in Arm A, but not in Arm B. The effect on iron status was observed when bLf is administered under fasting conditions, i.e., in the absence of active proteases. |
| **Infection** | | | |
| (Ajello et al., 2002) | Trial of 12 children with pharyngitis and scheduled for tonsillectomy given gargle with erythromycin and bLf | No adverse effects related to bLf administration were reported. | Combination erythromycin and bLf lowered the number of Group A streptococci compared with erythromycin alone. |
| (Berthon et al., 2022) | Systematic Review and Meta-analysis of Respiratory Tract Infections: 25 studies included with 10 examining evidence for effect on incidence, duration or severity of respiratory illness. | No adverse effects related to bLf administration were reported. | Lf was not associated with a reduction in respiratory tract infection incidence compared with control. Less than half of the included trials reported beneficial changes in immune cell phenotype or immune cell function. The available evidence indicates that a daily dose of 200 mg Lf may reduce IL-6 concentrations in some subject populations. However, due to the small number of trials and heterogeneous study designs, future research is required to determine effects on immune function and optimal supplementation strategies. |
| (Björmsjö et al., 2022) | Double-blind controlled trial, term formula-fed (FF) Swedish Infants (n = 180) randomized to receive bLf (1 g/L in a low iron formula) from 6 weeks to 6 months of age. | No adverse effects related to bLf administration were reported. | The study did not to show any evident or lasting  effect on inflammatory response or morbidity when lowering iron content or adding bovine lactoferrin to infant formula given to healthy term infants. No significant differences in otitis, respiratory infections, gastroenteritis, or other monitored infections and treatments were detected for any of the study feeding groups during the first 6 months and only a few and diverging effects were observed between 6 and 12 months |
| **(Giunta et al., 2012)** | **Prospective study of 21 pregnant women suffering from iron deficiency anemia, at risk of preterm delivery received 100 mg rhLf twice a day for one month, the other group received 520 mg ferrous sulfate** | **No adverse effects related to bLf administration were reported.** | **After 30 days, there was a significant reduction in abnormal vaginal flora in the LF group as compared to the ferrous sulfate group. There was also a decrease in IL-6 levels in bLf treated women. Pregnancy continued normally in both groups.** |
| **(Guntupalli et al., 2013)** | **Prospective, randomized, double-blind, placebo-controlled, multicenter phase 2 trial of 194 adults within 24 hours of the onset of severe sepsis; subjects given talactoferrin (1.5 g) or placebo every 8 hours for 28 days or until ICU discharge. Patients received standard of care for sepsis.** | **The drug was well tolerated with a safety profile like that of placebo.**  **(NOTE: Talactoferrin was evaluated globally in another phase 2/3 sepsis trial. The primary endpoint of this latter study was to determine the effect of oral talactoferrin alfa on 28-day all-cause mortality in patients with severe sepsis. Based on the review of available data, the DSMB for this second study recommended that patient enrollment and treatment be stopped).** | **The all-cause mortality at 28 days was 26.9% in the placebo group and 14.4% in the talactoferrin group (two-sided p = 0.052), representing a 12.5% absolute and a 46.5% relative reduction in mortality, meeting the protocol-specified primary endpoint. Reduction in all cause mortality was sustained at 6 months (p = 0.039).** |
| **(Laffan et al., 2011)** | **Randomized, double-blind study of thirty nursing-home residents randomized to receive, by gastrostomy tube, rhLf (5 mg/mL) or placebo via a flush solution. The solution was administered each day for 56 days. Nurses and nursing assistants recorded stool quality on each shift, and stool samples were tested for C. difficile at enrollment, day 14, 42, and 56. Efficacy in preventing post-antibiotic diarrhea was assessed.** | **Administering high concentrations (5 mg/mL) of rhLf produced no adverse effects in the population of frail older adults.** | **22 patients (13 control and 9 rhLf) completed the study. Fewer patients in the lactoferrin group experienced diarrhea compared to controls (statistically significant). There was no difference in C. difficile infection between groups. Caution should be taken in interpreting these results because this study was designed as a pilot project and enrolled a small number of patients. Further, the study protocol was altered mid-way through the project to achieve balance in treatment groups.** |
| (Mizuki et al., 2020) | Randomized, double-blinded, placebo-controlled parallel-group comparative trial. The eligible subjects were healthy adults working at kindergartens and nursery schools. Subjects randomized to the Placebo group (0 mg/day), the Low bLf group (200 mg/day), and the High bLf group (600 mg/day) for 12 weeks. | No adverse effects related to bLf administration were reported. | Prevalence of subjective acute gastrointestinal symptoms was significantly lower in the High bLf and the Low bLf groups than in the Placebo group. |
| (Ochoa et al., 2008) | Randomized, double-blind, placebo controlled, parallel group trial of children 12-36 month of age living in Peru given either 0.5 g bLf or 0.5 g maltodextrin (control) for nine months. Twenty-six children enrolled in each group; six children dropped out of each group. | No serious adverse events related to the intervention. | Comparison of overall diarrhea incidence and prevalence rates found no significant difference between the 2 groups. Colonization rates with common pathogens were similar in both groups, except for a statistically significantly lower frequency of Giardia positive samples in the bLf group. |
| (Ochoa et al., 2013) | Randomized double-blind placebo-controlled trial of 555 children in Peru (65 dropped out) comparing supplementation with bLf versus placebo. Previously weaned children were enrolled at 12–18 months and followed for 6 months. Children received 0.5g twice a day of bLF or maltodextrin placebo. | There were 17 severe adverse events (SAE), 9 bLF and 8 placebo. All SAE were hospitalizations for common pediatric illnesses; none were considered related to the intervention. | The diarrhea incidence was not different between groups. |
| (Ochoa et al., 2015) | A pilot randomized placebo-controlled double-blind study in 190 infants with a birth weight <2500g in three Neonatal Units in Peru. Patients were randomized to receive bLf 200mg/kg/day or placebo for four weeks. | There were no serious adverse events attributed to the intervention. | There was no statistically significant difference in frequency between groups of late-onset sepsis within 4 weeks from enrollment. |
| (Oda et al., 2021) | A randomized, double-blinded, placebo- controlled parallel-group comparative trial in healthy adults given placebo (n=99), 200 mg bLf (n= 95), or 600 mg/day of bLf (n=96) for 12 weeks. | Adverse events were similar among groups and were not related to test articles. | As a primary endpoint, no significant differences were observed in the prevalence of infectious diseases. The duration (days) of total infectious diseases was shorter in the 200 mg group and 600 mg group than in the placebo group. The duration of summer colds was shorter in the 600 mg group than in the placebo group. No changes in immune parameters were seen. |
| (Okada et al., 2002) | A dose escalation study was conducted in subjects with chronic hepatitis who received bLf tablets that were administered orally 2 or 3 times a day; 15 patients were entered at each of three dose levels (1.8, 3.6 and 7.2 g/day) for 8 weeks after which they were followed for the next 8 weeks. | bLf was well tolerated by subjects. | There was no effect of bLf on the primary endpoint of change in the serum ALT levels. There was no effect of bLf on secondary endpoints of changes in the serum hepatitis C virus RNA levels, virological (a 50% or greater decrease in hepatitis C virus RNA level) and biochemical (a 50% or greater decrease in the serum ALT level) responses at the end of treatment. |
| (Okuda et al., 2005) | Randomized, double-blind, placebo-controlled study of 59 subjects (34 adults and 25 children) positive for H. pylori infection. The bLf group received tablets of 200 mg b.i.d. for 12 weeks; the control group received placebo tablets. Subjects were assessed at the start of, end of and 4 weeks after end of administration using ^13^C-urea breath test (UBT). | No adverse effects related to bLf administration were reported. | A positive response (more than 50% decrease of the UBT value) was seen in 10/31 bLf treated subjects and 1/28 control subjects, suggesting that bLf administration reduced H. pylori colonization density (but did not lead to eradication). |
| (Sachdeva and Nagpal, 2009) | Meta Analysis of randomized controlled trials to determine the efficacy of bLf in H. pylori eradication. | No adverse effects related to bLf administration were reported. | Five RCTs involving 682 participants (316 in the experimental group and 366 in the control group) met the predefined inclusion criteria. There was potential improvement in H. pylori eradication. The methodological quality of the studies reviewed indicated that most studies were not of good quality. Three of the studies did not conceal allocation and in two studies it was inadequate or unclear. Four studies conducted intention-to-treat analysis. Only one study was double-blinded and placebo-controlled. The conclusions are based primarily on eradication data using author defined cutoffs, which were variable. Most of the trials (except one) did not specifically evaluate change in urea breath test values allowing only dichotomous evaluation of eradication. The interpretation of the results is also limited by the small number of trials available and the poor methodological quality of the trials (only one double-blind RCT). |
| **(Guttner et al., 2003)** | **Nine healthy subjects with minimal upper gastrointestinal symptoms and a positive urea breath test were recruited. None of the volunteers had previously been treated for H. pylori. Subjects received 5 x 1.0 g rhLf doses daily for 5 or 14 days. Breath tests were repeated during therapy and shortly after to check for eradication.** | **The safety and tolerability of the drug were assessed by physical examination and clinical laboratory evaluation (urea and electrolytes, full blood count, urinalysis, serum iron, ferritin, b-human chorionic gonadotropin (b-HCG)) before and after treatment, as well as monitoring of adverse events. No significant adverse effects were observed. No changes in routine laboratory results were observed.** | **No conversion of the urea breath test from positive to negative was observed and there was no consistent change in urea breath test count to indicate a possible suppression of H. pylori.** |
| **(Opekun et al., 1999)** | **Open label study in healthy but H. pylori infected subjects. The rhLf was administered by mouth five times throughout a 24-h period. Each dose of rhLf was mixed with 60 mL of whole milk until the rhLf was completely suspended. Six subjects received 250 mg per dose and six subjects 1 g per dose. Outcome was assessed by urea breath test.** | **No treatment related adverse events occurred.** | **rhLf was not effective in this study.** |
| (Tanaka et al., 1999) | Eleven patients with chronic hepatitis C received an 8-week course of bLf (1.8 or 3.6 g/day). | No adverse effects related to bLf administration were reported. | Alanine transaminase and hepatitis C virus (HCV) RNA concentrations decreased in 4 patients with low treatment serum concentrations of HCV RNA. In 7 patients with high pretreatment concentrations there was no change in these indices. |
| (Ueno et al., 2006) | Patients with chronic hepatitis C randomly received either oral bLF at a dose of 1.8 g daily for 12 weeks, or an oral placebo. The primary endpoint was the virologic response, defined as a 50% or greater decrease in serum HCV RNA level at 12 weeks compared with the baseline. The secondary endpoint was the biochemical response, which was defined as a 50% or greater decrease in the serum alanine aminotransferase (ALT) level at 12 weeks compared with the baseline. One hundred and ninety-eight of 199 patients were evaluable for efficacy and safety. | bLF treatment was well tolerated and no serious toxicities were observed. | There was no significant difference in virologic response rates between the two groups. In addition, bLF intake did not have any favorable effect on the serum ALT level. |
| (Yamauchi et al., 2000) | In a double-blind, randomized placebo-controlled study, doses of either 600 mg or 2000 mg bLf, or a placebo was orally administered daily for 8 weeks to 37 adults who were judged to have mild or moderate tinea pedis. | No adverse events and no subject withdrew from the study because of an adverse event. | A mycological cure was not seen in any of the subjects. |
| **COVID-19** | | | |
| (Algahtani et al., 2021) | Randomization into a control and two treatment groups ensured all groups received the approved Egyptian COVID-19 management protocol; only treatment group participants received bLf at different doses for seven days. | No adverse effects related to bLf administration were reported. | No statistically significant difference among studied groups regarding recovery of symptoms or laboratory improvement. |
| (Matino et al., 2023) | A total of 218 hospitalized adult patients with moderate-to-severe COVID-19 were randomized to receive 800 mg/day oral bLf (n = 113) or placebo (n = 105), both given in combination with standard COVID-19 therapy. | bLf showed an excellent safety and tolerability profile. | No differences in lactoferrin vs. placebo were observed in the primary outcomes: the proportion of death or intensive care unit admission or proportion of discharge or National Early Warning Score 2 within 14 days from enrollment. Results do not support its use in hospitalized patients with moderate-to-severe COVID-19. |
| (Navarro et al., 2023) | A randomized, double-blinded, placebo-controlled clinical trial was conducted in 209 participants from two tertiary hospitals that provide care to patients with SARS-CoV-2 infection in Lima, Peru. Daily supplementation with 600 mg of enteral bLF for 90 days was compared to placebo. | The numbers of subjects with adverse events were 49 in the bLF group and 32 in the placebo group. There was no significant difference among groups and none of the adverse events had a causal relationship with the intervention. | The study was prematurely cancelled due to the availability of vaccines against SARS-CoV-2 in Peru. SARSCoV- 2 infection occurred in 11 (10.6%) participants assigned to bLF and in 9 (8.6%) participants assigned to placebo without significant differences. There was no significant effect of bLF on time to symptomatic infection. |
| **Anti-inflammatory** | | | |
| (Chan et al., 2017) | Randomized, double-blind, placebo-controlled trial, 168 subjects aged 13–40 years old were randomly assigned to take either a capsule formulation containing 100 mg bLf with vitamin E and zinc or placebo twice a day for 3 months to assess number of acne lesions (inflammatory lesions). | No adverse effects related to bLf administration were reported. | The bLf group showed a significant median percent reduction in total lesions as early as 2 weeks with the maximum reduction occurring at week 10 compared to placebo group. Maximum reduction in comedones and inflammatory lesions also seen at week 10. |
| (Dix and Wright, 2018) | Double-blind randomized, cross-over trial was conducted with 12 healthy males randomized to one of two doses, equivalent to 200 mg or 600 mg lactoferrin, for two four-week supplementation arms, with a two-week washout period. Subjects received either standard bLf or Inferrin™ (microencapsulated bLf) for each arm to assess effect on immune markers and the microbiome. | No adverse effects related to bLf administration were reported. | The mean level of CD69+ activation on the CD4+ cells was lower after supplementation regardless of the form or dose of lactoferrin. Some subtle changes in the microbiome appeared such as decreased levels of Euryarchaeota, Acidobacteria, Chloroflexi, NC10,and Nitrospirae, and increased levels of Firmicutes and Bacteroidetes. |
| (Ishikado et al., 2010) | Liposomal-bLF composed of soy phosphatidylcholine was given as a supplement for four weeks in tablet form (180 mg bLF/d) to twelve subjects with periodontal disease with multiple sites of more than 3 mm probing depth (PD). PD, bleeding on probing (BOP), gingival crevicular fluid (GCF) volume and the levels of tumor necrosis factor (TNF)-a, interleukin (IL)-1b , IL-6, and monocyte chemoattractant protein-1 (MCP-1) in GCF were evaluated for 51 sites with more than 4 mm PD in five subjects. | No changes in blood chemistry parameters support safety of intake. | The PD was significantly reduced by L-bLF supplementation, but the BOP and GCF volume were not significantly changed. The MCP-1 level in GCF was significantly reduced, while levels of other cytokines were not changed. |
| (Mohamed et al., 2019) | An open-label, randomized, controlled pilot study; the effects of bLf on biochemical and behavioral markers in Alzheimer’s patients were evaluated in fifty patients (28 men and 22 women) with a clinical diagnosis of probable Alzheimer’s. | No adverse effects related to bLf administration were reported. | Improvement in Alzheimer’s surrogate markers of inflammation post treatment was reflected in enhanced cognitive function assessed by the Mini-Mental State Examination (MMSE) and Alzheimer's Disease Assessment Scale-Cognitive Subscale 11-item (ADAS-COG 11) questionnaires as clinical endpoints. Note: 3-month time period is too short for an Alzheimer’s study and the beginning MMSE scores of 19 suggest difficulty in completing subsequent tests. Transient improvements over short term can be due to the nature of the disease. |
| (Mueller et al., 2011) | An open-label, single-arm study, 43 adolescents and young adults were enrolled to take a chewable tablet formulation of bLf twice daily for 8 weeks. | None of the subjects experienced a bLf related adverse event during the trial. | At the end of the study (week 8), a mean reduction in inflammatory lesion count was not significant, significant reduction in noninflammatory lesion count of 23.5% and in total lesion count of 22.5% was observed as compared with baseline. |
| (Muscedere et al., 2018) | Phase 2, multicenter, randomized, double-blind, placebo controlled study of 212 (107 bLf and 105 placebo) patients expected to require invasive mechanical ventilation more than 48 hours received bLf both enterally and via an oral swab or a placebo of sterile water for up to 28 days. | There were no serious adverse events reported. | Antibiotic-free days and nosocomial infections did not differ between bLf and placebo groups. Clinical outcomes for lactoferrin versus placebo did not differ between groups: they were ICU length of stay, hospital length of stay, hospital mortality and 90-day mortality. Biomarker levels did not differ between the groups.  Conclusion: bLf did not improve the primary outcome of antibiotic-free days, nor any of the secondary outcomes. |
| **(Troost et al., 2003)** | **A randomized crossover dietary intervention of 15 healthy volunteers given a sucrose and a drink containing 5 g rhLf or placebo during breakfast. Subsequently, subjects ingested the same drink with 75mg of the NSAID indomethacin and after an overnight fast subjects consumed the drink and 50 mg indomethacin. A permeability test was performed to assess gastroduodenal and small intestine permeability as an indicator of NSAID-induced gastroenteropathy.** | **No side effects of rhLf ingestion were observed.** | **Small intestine permeability after indomethacin and placebo was significantly higher compared to the permeability observed after ingestion of indomethacin and rhLf. Gastroduodenal permeability did not differ between groups.** |
| **(Vincent et al., 2015)** | **A randomized, placebo-controlled, phase II/III clinical study of patients with severe sepsis receiving antimicrobial therapy.** **Patients were randomized to receive either talactoferrin (1.5 g, 15 mL) or placebo three times a day orally or by another enteral route for 28 days or until ICU discharge.** | **The study was terminated after 305 patients had been enrolled (153 talactoferrin and 152 placebo) because of futility and safety concerns identified by the Data Safety Monitoring Board.** | **Study terminated.** |
| **(Sortino et al., 2019)** | **A randomized, double-blind, crossover clinical trial investigated the effects of oral rhLf (1500 mg twice daily) versus placebo. The trial consisted of two 3-month treatment periods separated by a 2-4-month “washout” period. study of a rhLf treatment among persons with HIV, given in addition to antiviral therapy as a strategy to reduce systemic inflammation.** | **The most common adverse effect was gastrointestinal, but the frequencies were again similar in those receiving rhLf and those receiving placebo.** | **None of the inflammatory or immunologic measures assessed significantly declined with rhLf versus placebo treatment. No effects on the intestinal microbiome were seen.** |
| **Cancer** | | | |
| **(Hayes et al., 2006)** | **A Phase I trial with ten adult patients with progressive advanced solid tumors who had failed conventional chemotherapy were administered oral TALACTOFERRIN at doses from 1.5 to 9 g/day, using a 2 weeks on, 2 weeks off schedule. Patients were evaluated for drug toxicity, tumor growth rate, talactoferrin pharmacokinetics and cytokine markers.** | **Talactoferrin was very well tolerated. No hematological, hepatic, or renal toxicities were reported.** | **Significant levels of talactoferrin were undetectable in circulation, but a statistically significant increase in circulating IL-18, a pharmacodynamic indicator of talactoferrin activity, was observed** |
| **(Hayes et al., 2010)** | **A phase IB study to evaluate the safety and activity of oral talactoferrin in patients for whom no effective therapies were available. Trial included 36 patients; dose escalation phase of the trial used doses of 1.5, 4.5, or 9 g/day. Second part of the study patients were randomly assigned to receive one of the two highest doses (4.5 or 9 g/day) administered in two divided doses.** | **Talactoferrin was very well tolerated with no test article related SAE or grade ¾ adverse effects either clinically or in laboratory findings. There were no detectable hematological, renal, or hepatic toxicities, and no apparent dose-dependence to any of the toxicities.** | **The growth rate of prospectively defined target lesions decreased substantially (14% absolute and 70% relative reduction; P<0.01).** |
| (Iigo et al., 2014; Kozu et al., 2009) | Blinded, randomized controlled trial. Patients with colorectal polyps ≤5 mm diameter and likely to be adenomas ingested 0, 1.5, or 3.0 g bLF daily for 1 year. | No adverse effects related to bLf administration were reported. | The authors concluded that bLF did not induce an immune response against the polyps. Nineteen participants had target polyps that had increased in size by 20 %, 11 participants had target polyps that had decreased in size by 20 % or more, and the remaining 73 participants had polyps that were diagnosed as having no change in size. Trial participants with regressing polyps had increased NK cell activity, increased serum hLF levels (indicating increased neutrophil activity), and increased numbers of CD4+ cells in the polyps. The colorectal polyps examined persisted throughout the one-year trial period in the patients ingesting 3 g bLF daily, indicating that the polyp-associated CD4+ and NK did not induce an immune response against the polyps. This is also consistent with the suggestion that ingestion of bLF primed rather than activated immune effector cells. This premise would predict that ingestion of bLF would not change immune function from OFF to ON, but rather immune function would change from a less responsive state to a more responsive state. Participants 63 years old and younger ingesting 3.0 g bLF daily for one year had a statistically significant regression in the growth of colorectal polyps compared with participants ingesting placebo, however, bLF had no significant effect on polyps in participants 64 years old and older. |
| **(Jonasch et al., 2008)** | **Forty-four adult patients with progressive advanced or metastatic RCC who had failed prior systemic therapy received oral talactoferrin at a dose of 1.5 g twice daily on a 12-week-on 2-week-off schedule. Patients were evaluated for progression-free survival at 14 weeks, overall response rate, and progression free and overall survival.** | **Talactoferrin was well tolerated. No significant hematologic, hepatic, or renal toxicities were reported.** | **In this preliminary study, the study met its predefined target with a 14-week progression-free survival rate of 59%. The response rate was 4.5%. The median progression-free survival was 6.4 months and the median overall survival was 21.1 months.** |
| **(Parikh et al., 2011)** | **Patients (n = 100) with stages IIIB to IV non-small-cell lung cancer for whom one or two prior lines of systemic anticancer therapy had failed were randomly assigned to receive either oral talactoferrin (1.5 g in 15 mL phosphate-based buffer) or placebo (15 mL phosphate-based buffer) twice per day in addition to supportive care. Oral talactoferrin or placebo was administered for a maximum of three 14-week cycles with dosing for 12 consecutive weeks followed by 2 weeks off. The primary objective was overall survival (OS) in the intent-to-treat (ITT) patient population. Secondary objectives included progression-free survival, disease control rate and safety.** | **Talactoferrin was well tolerated and, generally, there were fewer adverse events (AEs) and grade ≥ 3 AEs reported in the talactoferrin arm.** | **Talactoferrin was associated with statistically significant improvement in OS in the ITT patient population.** |
| **(Ramalingam et al., 2013)** | **Phase III, randomized, double-blind, placebo controlled trial of talactoferrin in patients with histologically or cytologically confirmed stage IIIB/IV non-small-cell lung cancer whose disease had failed two or more prior systemic anti-cancer regimens, including at least one platinum-containing regimen, for advanced or metastatic IV non-small-cell lung cancer. Patients were randomly assigned in a 2:1 ratio (497:245 patients) to receive oral talactoferrin at a dose of 1.5 g b.i.d. or placebo b.i.d., respectively. Treatment was administered for a maximum of five 14-week cycles. The primary efficacy end point was overall survival (OS); secondary end points included 6- and 12-month survival, progression-free survival and disease control rate.** | **Safety profiles were comparable between arms.** | **Talactoferrin did not produce any improvement in efficacy as measured by the primary and secondary endpoints.** |
| **(Digumarti et al., 2011)** | **Double-blind, randomized, placebo-controlled trial; patients were randomly assigned (1:1) to one of two treatment arms without stratification:**  **Arm 1: Carboplatin + paclitaxel (175 mg/m2); C/P + oral talactoferrin (1.5 g in 15 ml twice daily for three 6-week cycles).**  **Arm 2: C/P at the same doses as in arm 1 + oral placebo (15 ml twice daily for three 6-week cycles).** | **Adverse events were consistent with chemotherapy.** | **The trial met the primary end point of improvement in confirmed response rate in the prospectively defined evaluable population. Compared with the placebo group, response rate increased in the talactoferrin group by 18% and 15% in the evaluable and intent-to-treat populations, respectively.** |
| **Anti-Obesity** | | | |
| (Ono et al., 2010) | Double-blind, placebo-controlled study, Japanese men and women (n = 26; aged 22–60 years) with abdominal obesity (BMI > 25 kg/m2, and visceral fat area (VFA) > 100 cm2) consumed enteric coated bLf (300 mg/d) or placebo tablets for 8 weeks. | No adverse effects of the bLf treatment were found with regard to blood lipid or biochemical parameters. | Significant decreases in body weight, BMI, hip circumference, total fat area, visceral fat area and subcutaneous fat area were noted in the treated group compared to placebo. |
| **Neonates NEC** | | | |
| (Akin et al., 2014) | Prospective, placebo-controlled, double-blind, randomized trial, infants either VLBW or born before 32 weeks received bLf (200 mg/day) or placebo (n=25 in each group). | No adverse effects related to bLf administration were reported. | Fewer nosocomial sepsis episodes were observed in bLf treated infants (statistically significant). |
| **(Sherman et al., 2016)** | **A randomized, double blind, placebo-controlled trial in 60 infants per group with birth weights of 750 to 1500 grams was conducted. Each infant received enteral talactoferrin or placebo on day 1 through 28 days of life; talactoferrin dose was 150 mg/kg/12 hour. The original power calculations specified a minimum of 360 subjects but due to reduced funding, only 120 infants were randomized. Primary outcomes were bacteremia, pneumonia, urinary tract infection, meningitis, and necrotizing enterocolitis. Secondary outcomes were sepsis syndrome and suspected NEC.** | **No treatment related adverse events occurred. Rates of SAE were also similar between treatment and placebo arms. No differences in growth or neurodevelopment occurred among infants treated with talactoferrin and placebo during a one-year, post-hospitalization period.** | **Because of the reduction in sample size the investigation was underpowered to identify significant primary or secondary outcomes. Hospital-acquired infections in the group with talactoferrin were 50% of that observed in infants fed placebo (p<0.04), including fewer blood or line infections, urinary tract infections, and pneumonia.** |
| (Ali et al., 2021) | Meta-analysis of 6 randomized controlled trials of bLf supplemented infant formula or capsules/tablets | No adverse effects related to bLf administration were reported. | Significantly reduced odds of the development of respiratory tract infections with the use of bLf relative to regular formula or placebo. |
| (Barrington et al., 2016) | Pilot trial of very preterm infant (n = 79) receiving 100 mg/day of bLf for control. The primary outcome was feeding tolerance. | No adverse effects reported; bLf well tolerated. | Mortality, late onset sepsis and other complications of prematurity were no different between treated and control groups. |
| (Dobryk et al., 2022) | Prospective cohort study included 126 preterm infants with a gestational age of < 32 weeks, a birth weight of < 1,500 g, | No adverse effects related to bLf administration were reported. | Enteral supplementation with bLF at a dose of 100 mg/day did not reduce the incidence of LOS, NEC, ROP, severe CNS lesions, and overall mortality in preterm very low birth weight infants |
| (Kaur and Gathwala, 2015) | Randomized, double-blind, placebo-controlled trial with asymptomatic neonates, <2000 g, admitted to NICU in first 12 h of birth with no maternal risk factors for sepsis were randomized to receive bLf or placebo from 1st to 28th day of life. 130 infants received bLf and 67 received placebo. Doses based on body weight ranged from 80-142 mg/kg/day mixed in milk. | In a 1-month follow-up period, there were no apparent adverse effects recorded. No participant required discontinuation owing to intolerance. Neonates who received bLf showed a significantly better tolerability towards feeds. | The incidence of the first episode of culture-proven late-onset sepsis was significantly lower in the bLf group vs. placebo. Statistically significant reduction in the sepsis-attributable mortality was also seen after use of prophylactic bLf. The incidence of bacterial and fungal sepsis was not statistically significantly different between groups. |
| (Manzoni et al., 2014) | Randomized, double-blind, placebo-controlled trial of 743 neonates assessed until discharge for development of NEC. Infants were randomly assigned to receive orally either bLf (100 mg/day) alone (group LF; n = 247) or with LGG (at 6×109 CFU/day; group bLf + LGG; n = 238), or placebo (Control group; n = 258) from birth until day 30 of life (45 for neonates <1000 g at birth). | No adverse effects or intolerances to treatment occurred. | The incidence of death-and/or-NEC was significantly lower in both treatment groups vs control. |
| (Griffiths et al., 2018) | ELFIN trial: Large multicenter randomized, double-blind, placebo- controlled parallel group prospective trial of very low birth weight neonates (weight less than 1500 g, aged less than 8 days) born before 32 weeks’ gestation in 37 UK hospitals and 97 continuing care sites until 34 weeks’ postmenstrual age. The bLF was administered once daily by nasogastric or orogastric tube or orally once the enteral feed volume was > 12 ml/kg/day and continued until 34 weeks’ postmenstrual age. Dose of bLf was 150 mg/kg per day; maximum 300 mg/day; (n = 1098) or sucrose (control group) (n = 1101). Some small infants may have had the dose split at the discretion of the responsible clinical team. A maximum of 70 days of treatment was given. | Adherence to the intervention was high, the proportion of protocol violations was low, and outcome data were available for more than 99% of the cohort. Few adverse or intolerance events related to the intervention were observed. | Primary outcome was microbiologically confirmed or clinically suspected late-onset infection:  bLF (150 mg/kg per day until 34 weeks’ postmenstrual age) does not reduce the risk of late-onset infection, other morbidity, or mortality in very preterm infants.  There were no differences between groups in the risk of morbidity or on levels of care received; there were no reported effects on the degree of exposure to antimicrobial drugs, on the duration of hospitalization, or on stay in intensive care settings.  The conclusion from this study is that supplementation with bLF does not reduce the incidence of infection, mortality, or other morbidity in very preterm infants. |
| (Embleton et al., 2021) | Longitudinal impact of lactoferrin in healthy preterm infants in 13 NICU sites; to explore the actions of lactoferrin affecting the gut, a subset of infants from the ELFIN trial (Mechanisms Affecting the Gut of Preterm Infants in Enteral Feeding (MAGPIE) study) was identified who did not develop confirmed necrotizing enterocolitis or late-onset sepsis and matched with samples from healthy control infants. Very preterm infants born before 32 weeks’ gestation in 37 UK hospitals and 97 continuing care sites until 34 weeks’ postmenstrual age. Subjects were younger than 72 h at randomization.  479 preterm infants were recruited and collected  > 33,000 usable samples from 467 infants. 16S ribonucleic acid bacterial analysis was conducted on samples from 201 infants, of whom 20 had necrotizing enterocolitis and 51 had late-onset sepsis, along with samples from healthy matched controls to explore longitudinal changes. | Nested within the ELFIN RCT | Although lactoferrin significantly decreased the level of Staphylococcus and other key species, the impact was much smaller than that of other clinical variables, such as infant age or hospital site. This is in keeping with the results of the ELFIN trial, which showed no reduction in late-onset sepsis or NEC. Investigators observed minimal, if any, impact of lactoferrin on the metabolome. |
| (Young et al., 2023) | Parents of preterm infants <32 weeks’ gestation who were enrolled in the ELFIN provided signed consent to collect stool and urine from their baby. 467 usable samples totaling 10,990 stool and 22,341 urine samples. Analyses of gut microbiome (1304 stools, 201 infants), metabolites (171 stools, 83 infants; 225 urines, 90 infants) and volatile organic compounds (314 stools, 117 infants) were performed. | Nested within the ELFIN RCT. | Gut microbiome Shannon diversity at 34 weeks corrected age was not significantly different between infants in the lactoferrin or placebo groups. This multiomic study identified minimal impacts of lactoferrin but much larger impacts of hospital site and postnatal age. This may be due to the specific lactoferrin product used, but more likely supports the findings of the RCT in which this study was nested, which showed no impact of lactoferrin on reducing rates of sepsis. |
| *Studies in bold are done with rhLf, non-bolded studies used bLf. Review does not include studies where lactoferrin was evaluated in combination with another test article or part of another product such as whey protein isolate. Studies were included that administered lactoferrin along with standard of care. Clinical studies by oral administration only were reviewed. | | | |

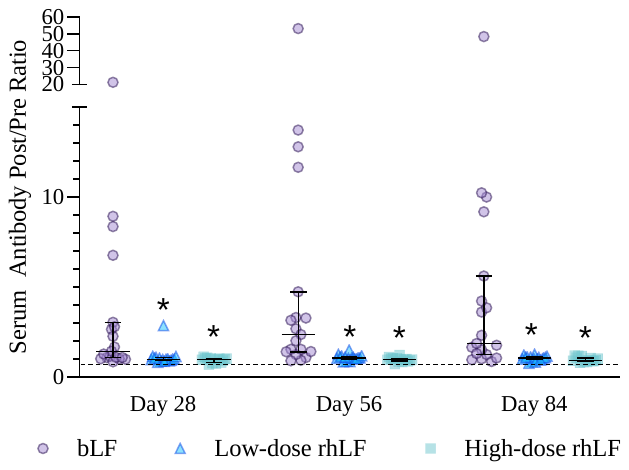

**Supplemental Figure 1.** Median, interquartile limit ranges, and individual participant responses for the change from baseline in serum anti-lactoferrin antibody (expressed as the post/pre ratio) at Days 28, 56, and 84 in the bLF, low-dose rhLF, and high-dose rhLF groups in the per protocol population of Study 1. The horizontal dashed gray line represents a post/pre ratio of 2, a conservative threshold above which the presence of study product-emergent antibodies is unlikely due to random variability. * Indicates that the change in serum anti-LF antibodies from baseline to the respective day was significantly different from the bLF group (P < 0.01). Abbreviations: bLF, bovine lactoferrin; rhLF, human recombinant lactoferrin.
